# Modeling and Multiobjective Optimization of Indoor Airborne Disease Transmission Risk and Associated Energy Consumption for Building HVAC Systems

**DOI:** 10.1101/2021.08.10.21261866

**Authors:** Michael J. Risbeck, Martin Z. Bazant, Zhanhong Jiang, Young M. Lee, Kirk H. Drees, Jonathan D. Douglas

## Abstract

The COVID-19 pandemic has renewed interest in assessing how the operation of HVAC systems influences the risk of airborne disease transmission in buildings. Various processes, such as ventilation and filtration, have been shown to reduce the probability of disease spread by removing or deactivating exhaled aerosols that potentially contain infectious material. However, such qualitative recommendations fail to specify *how much* of these or other disinfection techniques are needed to achieve acceptable risk levels in a particular space. An additional complication is that application of these techniques inevitably increases energy costs, the magnitude of which can vary significantly based on local weather. Moreover, the operational flexibility available to the HVAC system may be inherently limited by equipment capacities and occupant comfort requirements. Given this knowledge gap, we propose a set of dynamical models that can be used to estimate airborne transmission risk and energy consumption for building HVAC systems based on controller setpoints and a forecast of weather conditions. By combining physics-based material balances with phenomenological models of the HVAC control system, it is possible to predict time-varying airflows and other HVAC variables, which are then used to calculate key metrics. Through a variety of examples involving real and simulated commercial buildings, we show that our models can be used for monitoring purposes by applying them directly to transient building data as operated, or they may be embedded within a multi-objective optimization framework to evaluate the tradeoff between infection risk and energy consumption. By combining these applications, building managers can determine which spaces are in need of infection risk reduction and how to provide that reduction at the lowest energy cost. The key finding is that both the baseline infection risk and the most energy-efficient disinfection strategy can vary significantly from space to space and depend sensitively on the weather, thus underscoring the importance of the quantitative predictions provided by the models.

## 1. Introduction

With the recognition that airborne exchange of respiratory aerosols may be the primary means of transmission for COVID-19 and other diseases [1–4], it is crucial to understand how HVAC systems can affect this infection route [5]. In order to reduce the aerosol concentration and associated infection risk in buildings, HVAC systems can deliver clean air flow via a combination of high-efficiency filtration, extra fresh-air ventilation, and other sources. Unfortunately, each option leads to increased energy use, and their effect on indoor-air quality can vary significantly based on outdoor-air conditions. In this paper, we propose a model-based optimization framework to determine the combination of clean-air sources that can achieve a given reduction in infection risk in the most cost-effective manner. The optimization can recommend a spectrum of possible operational profiles ranging from minimum infection risk to minimum energy, providing predicted energy consumption and infection risk for each. This information can empower building managers to make informed decisions about how to operate their buildings given current cost and disinfection priorities.

Underpinning the proposed optimization is a set of dynamic models for zone temperature, humidity, and the expected transmission rate, which are parameterized in terms of physical and usage characteristics of the zone (e.g., floor area, ceiling height, design occupancy, HVAC equipment sizes) that are representative of typical commercial buildings. These models can then be used to predict the effect of various design and operational variables on energy and infection risk, thus allowing actionable and informed recommendations to be made. We begin by reviewing the relevant background for airborne transmission modeling in buildings along with the associated energy analysis, and then we present the main goals of this paper.

### 1.1. Airborne Disease Transmission

For airborne diseases, the primary transmission route is via viruses or bacteria that are entrained in aerosolized particles released into the air by exhalation of an actively infectious individual [3, 4]. These expiratory aerosols are partially dried droplets of respiratory fluids, such as saliva [6] or airway mucus [7], and they are composed of aqueous solutions of proteins and salts, along with potentially infectious microbes. The concentration and size distribution of the particles covers several orders of magnitude [8, 9], with significant dependence on vocalization volume [2, 10] and respiratory mode [11, 12]. The largest of these droplets, produced primarily by coughs and sneezes [13] can be up to 1 mm in diameter. However, because they settle rapidly to the ground, these large droplets generally do not pose a risk for long-range airborne transmission [14, 15]. By contrast, the smallest droplets are sized between 0.1 and 10 µm and are produced in high concentrations by standard respiration. Because these particles settle much more slowly, they tend to remain suspended in the air for long periods of time that exceed the time scales for turbulent mixing and air change in an indoor space [4]. Such aerosol particles are of primary concern for airborne transmission, as they can spread over long distances due to the air circulation provided by natural convection and HVAC systems [16–18], thus leading to new infections if inhaled by susceptible individuals.

Although infectious aerosols can remain airborne for long periods, their concentration does decay over time due to a variety of removal mechanisms [4]. For example, the particles naturally deposit onto surfaces (with a size-dependent rate [19, 20]) as fomites, thus rendering them unlikely to transmit the disease [21]. In addition, the particles may simply be removed from the air via outdoor-air ventilation (with particle-laden air exhausted to the ambient and replaced with a similar volume of outdoor air free from respiratory particles) or filtration (by which some fraction of the particles become trapped in a filter) [22–24]. In addition, the infectious material in the particles can become deactivated, e.g., due to natural decay (the rate of which depends slightly on temperature and humidity) or deliberate UV irradiation [25–28]. Thus, these mechanisms result in a gradual decay of the airborne infectious particle concentration over time. Ultimately, the concentration of infectious aerosols in the space is determined by the balance of generation and removal mechanisms. However, because conditions are often unsteady throughout the course of the day (e.g., with generation rates varying in accordance with the occupancy cycle and removal rates determined by the transient operation of the HVAC system), it is important to account for dynamic effects when considering infection risk.

Due to the significant likelihood and associated negative consequences of indoor disease transmission, there is growing interest in mitigating the associated risk by changing the design or operation of building HVAC systems. Many such strategies are aimed at reducing the generation rate of infectious particles within the space and reducing the exposure of susceptible individuals to those particles. For example, by requiring occupants to wear face masks, the particle concentrations released by exhaling infectors and received by inhaling susceptibles are reduced in accordance with the filtration efficiency of the mask [4, 29]. Masks also largely eliminate the momentum of respiratory flows, such as turbulent plumes from speech [30] or coughs and sneezes [14], which would otherwise enhance the risk of short-range airborne transmission [31] relative to the long-range airborne transmission in a well mixed space [4]. Furthermore, screening occupants for high temperatures or other symptoms prior to entering a building can significantly reduce the expected number of infectors in the space (and thus also the infectious particle generation rate), although the effectiveness of this strategy is attenuated for diseases such as COVID-19 for which asymptomatic spread is prevalent [6, 32]. By contrast, other commonly employed measures may have little to no effect on airborne transmission. For example, the installation of plexiglass dividers and enforcement of social distancing can reduce the likelihood of short-range transmission associated with respiratory jets [4, 6, 31] or large droplets [14, 15], but once the smaller particles have mixed with the surrounding air, these strategies no longer provide protection to susceptible occupants. These observations indicate that focusing on the long-range airborne transmission route is a more effective means of reducing infection rates. Indeed, recent analyses of COVID-19 in schools found no significant change in transmission rates resulting from 3-foot versus 6-foot social distancing requirements [33] or whether physical barriers were installed to separate desks, but reduced transmission was observed from mask usage, increased ventilation, and improved filtration [34].

Although these findings indicate possible strategies for reducing airborne transmission, and even which among them are likely to be most effective, the value of such *qualitative* guidance is somewhat limited. Specifically, knowing that a particular action provides *relative* reduction in infection risk does not indicate whether it is necessary in an *absolute* sense. Given the significant diversity of building operation and occupant behavior, the expected rate of indoor infection can vary over several orders of magnitude for different buildings or spaces [4]. Thus, in order to guide decision-making, it is important to provide *quantitative* guidance for various strategies.

#### 1.1.1. Modeling Framework

Mathematical models to estimate the rate of airborne disease transmission have been known for over 50 years, with seminal contributions from Wells [22] and Riley et al. [35]. Within the Wells-Riley framework, the hypothetical concentration of infectious aerosols is modeled using material balances, from which the received dose and infection probability of susceptible individuals in the space can be calculated. This basic approach has been extended in many directions since its original proposal [4, 19, 20, 23, 24, 36–41]. In general terms, the key relationships for one susceptible individual can be expressed as follows:

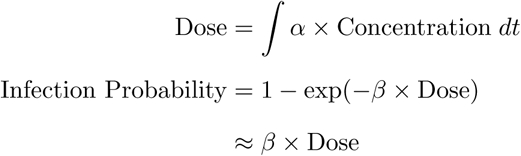

That is, the individual’s received dose (measured in some unit of amount, e.g., number or mass of virions) is equal to the time integral of the airborne concentration of infections particles (as modeled by the material balance) multiplied by an exposure rate *α* (generally equal to the individual’s breathing rate). That individual’s probability of infection is then given by an exponential relaxation to certainty proportional to the total dose, in which the proportionality constant *β* reflects the infectivity or hazard rate of the dose. From Wells [22], it is common to measure each amount of infectious particles in units of “infectious quanta”, where one “quantum” (abbreviated q) represents the amount of pathogen exposure that results in an infection probability of 1 − exp(−1) ≈ 0.63. (In these units, the proportionality constant *β* thus has a value of 1 q^−1^.). More generally, infection quanta may be defined in terms of the mean rate of transmission between each infected-susceptible pair [4], without regard to the microscopic pathogen concentration.

In order to determine the resulting infectious particle concentration, a common assumption is to assume that the particles are well-mixed (i.e., spatially uniform in concentration) within the air. Indeed, the same assumption is commonly made when modeling other mass and energy balances within buildings, and it can be justified both theoretically [4] and empirically via simulation [18] and experiment. Where the effects are relevant, the well-mixed models can be corrected to account for short-range respiratory jets [6, 31] or fluctuations in airflow [16], but it is nevertheless a useful first approximation to assume well mixed indoor air. Another complication is particle-size dependence of various processes, primarily the filtration efficiency and deposition rate, but also infectivity and decay rate [4]. These effects can be accounted for, but it is common to include them only via size-averaged variables in existing models [4, 20, 42]. Additional non-uniformity can result when a space is separated into multiple rooms that share the same supply air stream, but modeling in such cases is straightforward, if the room dimensions, occupancy, and airflow are known [43].

#### 1.1.2. HVAC Mitigation Strategies

As alluded to above, the operation of building HVAC systems can have a significant impact on the airborne concentration of infectious particles. The primary sources of removal and deactivation are ventilation, filtration, and UV disinfection. Some past studies have assessed which HVAC strategies are most effective and energy-efficient for infection risk reduction [37, 40], but findings can vary significantly based on climate and HVAC system design. A key focus from ASHRAE [44] and others has been on increasing ventilation rates. Indeed, increased ventilation does reduce the effective concentration of infectious particles, and in addition the associated reduction in indoor CO_2_ concentration is beneficial for general health and wellness [45]. However, if the additional ventilation is provided only by increasing the outdoor air *fraction* in the supply air stream, the supplemental reduction in infectious particle concentration can be small to negligible if that stream also passes through a filter with a high MERV rating [44]. Similarly, switching to a higher-efficiency filter will remove more infectious particles from the air, but if the volume of air passing through that filter is low, then absolute benefits may not be significant [37]. Thus, it is important to consider all the disinfection strategies available and account for how they interact when making decisions. In particular, to provide meaningful reduction in infection risk, it is often necessary to increase the *total* airflow provided by the HVAC system so as to more rapidly dilute and remove infectious particles present in the air.

A key caveat when considering HVAC infection mitigation strategies is that due to system design, it is often not possible to directly manipulate the desired variables. For example, a practitioner may want to double the ventilation and total airflow provided to a zone. However, in constant-volume systems, the supply flow generally cannot be increased above its design value, and so the only way to increase total flow is to install standalone filtration devices that operate on their own dedicated stream of air. Alternatively, in variable-volume systems, the flow delivered to a given room is determined by the action of the VAV controller for the purposes of temperature regulation, and so absolute flow can only be adjusted indirectly (e.g., by manipulating supply-air temperatures). Finally, if the system does not have an airside economizer, it may not be possible to increase ventilation rates above their minimum values, and even when present, the maximum ventilation rate may be further restricted based on weather conditions and heating/cooling capacities. The main observation is that when considering HVAC mitigation strategies for airborne infection risk, it is not sufficient to consider just the fundamental physical variables, and instead one must step further back to use the setpoints exposed by the HVAC system and predict their effect on the relevant physical variables.

### 1.2. Energy Consumption

Although HVAC systems can be operated so as to reduce the risk of airborne transmission of disease, unfortunately the actions taken will almost always *increase* energy consumption relative to baseline operation. Thus, when choosing between two strategies, it is important to consider which is most energy-efficient. A common basis to quantify the disinfection efficacy of a given mechanism is its removal rate of infectious particles, which is best expressed as the equivalent volumetric flow of particle-free air that would provide the same removal rate. This concept is referred to as the “clean air delivery rate” [46] or the “equivalent outdoor air” rate [44], and two devices that have the same value are thus equivalent from an infection-risk perspective and can be compared in terms of energy consumption. However, when combining multiple clean-air mechanisms in series (e.g., using an air-filter downstream from the outdoor-air intake), it is important to consider their *combined* efficacy, as there may be only incremental benefits to clean-air delivery but a significant change in energy consumption.

Given these associated energy costs and the societal emphasis on energy efficiency and sustainability, it is important to make informed decisions about which strategies to apply. This sentiment has been echoed by the ASHRAE Epidemic Task Force [47], which has recommended “select[ing] control options, including standalone filters and air cleaners, that provide desired exposure reduction while minimizing associated energy penalties”. In order to make this determination, it is necessary to estimate the energy consumption associated with possible courses of action. This knowledge will enable building managers to make informed design decisions while adapting operational decisions regularly to account for changing weather or infection-prevalence conditions.

#### 1.2.1. Filtration and Fan Power

A key technology for removing infectious particles from a space is filtration. The overall clean-air delivery rate associated with a filter is the product of the volumetric flow of air through the filter multiplied by its filtration efficiency [44]. This observation suggests two different ways to utilize filters, which have different impacts on operation and energy consumption.

The first way to increase the clean-air delivery provided by a filter is to increase the amount of air flowing through it. Physically, this increased flow is provided by increasing fan speed, which thus requires additional electricity consumption that can be calculated as the product of volumetric flow and pressure rise divided by overall fan efficiency. Because the pressure rise and efficiency each vary with flow, the result is essentially a static nonlinear model in terms of flow [48]. The simulation package EnergyPlus [49] supports various mathematical forms for variable-speed fan energy models, including low-order polynomials and a power law. Although model accuracy is best when calibrated against actual operating data, reasonable accuracy can be achieved by scaling standard models with nameplate design flow and power consumption of the fans [48].

As an alternative to increasing total flow, particle removal rates can be increased by switching to a filter with higher efficiency. The MERV rating standard specifies required filtration efficiencies for three different ranges of particle sizes, and thus for a given particle size distribution, these values can be used to calculate effective the overall filtration efficiency of each MERV rating [44]. Unfortunately, this increase in efficiency restricts the flow of air through the filter, thus increasing the required pressure difference to produce the same total flow rate. In general, the higher pressure difference results in higher energy consumption, although the magnitude can vary based on the fan control strategy that is applied [50]. To a reasonable approximation, the increased energy consumption can be estimated by considering the change in pressure drop associated with the new filter type and appropriately scaling the baseline energy consumption [37].

#### 1.2.2. Ventilation and Coil Loads

When supplying additional ventilation, there is a change the heating or cooling load necessary to maintain indoor air temperature, which thus results in a change in energy consumption. Past studies have quantified the extra costs associated with increased ventilation rates [45, 51, 52], and a key finding is that there is significant variation with climate. The impact of day-to-day variation can perhaps be minimized via the use of an economizer (which will automatically increase ventilation rates when outdoor conditions make it energetically favorable), but the commonly employed rule-based strategies are often insufficient to to capture all the available benefits [53]. In Azimi and Stephens [37], the energy costs of extra ventilation were explicitly compared to that of higher-efficiency filtration for various climates and filter types, finding that filtration is generally the more energy-efficient option overall. However, results can change seasonally with weather conditions, and the study did not consider the energy consumption associated with dehumidification. In general, it is recognized that ventilation is a costly component of HVAC system operation, and so [54] specifies *minimum* ventilation rates that must be provided and allows technologies like demand-controlled ventilation to further reduce the amount of ventilation that must be supplied. Energy-recovery technologies can be applied to reduce the energy impact of ventilation [55], but they are not commonly employed in existing buildings.

The energy costs associated with ventilation ultimately derive from the fact that the outdoor air may have to be heated or cooled to bring it to suitable conditions of temperature and humidity for indoor comfort. Thus, the energy consumption should be modeled by considering the thermal loads at the appropriate coils. For cooling coils, effectiveness models can capture heat-transfer limitations and thus provide an accurate estimate of energy consumption [56–58]. However, simplified contact-mixture models can also be used and have been shown to match experimental data despite their relative simplicity [59]. For heating coils, the modeling principles are similar but generally simpler because there is no associated change in humidity that must be considered as a latent load. The calculated coil loads can then be converted to the appropriate energy source (generally electricity or gas) via the appropriate coefficient of performance or efficiency factor [37].

The key differentiator between ventilation and filtration for the purposes of reducing airborne transmission is that for a given level of airflow or particle removal, the energy cost of filtration is essentially constant, whereas the energy cost of ventilation varies significantly (and can even become negative) based on outdoor-air conditions. Thus, optimal energy efficiency requires operating strategies to be adjusted seasonally (or more frequently), perhaps with filtration preferred in the extreme heating and cooling months and ventilation used during milder transition seasons.

#### 1.2.3. Other Disinfection Devices

For completeness, we note that infectious particles can also be removed or deactivated by auxiliary disinfection devices, including UV irradiation zones in ducts or standalone filters and air cleaners that are placed in the space. For UV disinfection, the radiation intensity to achieve a given level of disinfection can be computed from expected flow rates [28]. However, once the UV lamps have been installed, their electricity consumption is essentially constant. Thus, for energy analysis, it is only necessary to know the total rated power of the UV lamps (and of course also whether they are on or off). For filtration devices, theoretical models have been developed to estimate and optimize overall particle removal rate for a given electricity consumption [60], but as before, the main value relevant for energy analysis is the device’s rated power. Where variable operation is possible (e.g., for a device that can operate at different fan speeds or flow rates), it may be necessary to fit simple models to account for variation, possibly with data obtained from manufacturer specifications.

### 1.3. Overview of Paper and Contributions

Given the importance of managing both indoor infection risk and HVAC energy consumption [47], the goal of this work is to present dynamic models that can be used for monitoring and optimization of both variables in buildings. The models for indoor temperature, humidity, and hypothetical infectious particle concentration are based on mass and energy balances that follow from standard physics. By contrast, the model of the regulatory control system is phenomenological so as to match general HVAC system behavior (including time variation in airflow resulting from variation in thermal loads) while remaining computationally efficient. Both sets of models are parameterized in terms of readily available data, including space dimensions, equipment capacities, weather conditions, and occupancy profiles. Where values are not specifically known, they can be substituted with standard values provided in ASHRAE standards [54, 55]. The main contribution is that the models are coupled, and thus they predict simultaneously the infection rate, energy consumption, and indoor conditions that result from commonly available HVAC setpoints and design variables. Thus, these models can provide *actionable* guidance that can be readily applied in real buildings. In particular, they facilitate Pareto analysis to assess the optimal tradeoff between the two primary metrics so that both aspects can be considered.

The remainder of this paper is structured as follows. In Section 2, we present the full set of dynamic models for airborne infection, thermal dynamics, and the regulatory control system. For brevity, we present only the models themselves (i.e., equations and descriptions of variables), but additional detailed discussion is provided in the appendices. In Section 3, we then discuss how the models can be used for monitoring and optimization purposes. These calculations could be used by building managers to evaluate the infection risk in their buildings as operated and determine how infection rate could be reduced or energy efficiency can be improved. We then present examples in Section 4 to illustrate the insights and operational benefits that can be provided by application of the modeling framework. Finally, Section 5 provides conclusions and a discussion of future directions.

## 2. Modeling

In this section, we present various dynamic models that can be used to estimate and predict infection risk and HVAC energy consumption within an indoor zone. All models are defined in the standard continuous-time state-space form

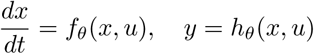

for states *x*, inputs *u*, outputs *y*, and static parameters *θ*. (Note the distinction between *u* and *θ* is that values of *u* vary throughout the simulation period, whereas values of *θ* are held constant.) Models are simulated using the standard 4th order explicit Runge-Kutta method assuming a piecewise-constant hold on *u* and calculating *y* as the *average* value over the integration interval, which for our purposes is Δ = 15 min.

Performance metrics are all calculated for each timestep as functions *ℓ*_*θ*_(*y, u*) that are then integrated as appropriate over the simulation interval. In various contexts, they are used as optimization objectives or constraints.

### 2.1. Airborne Disease Transmission

In order to predict the transmission risk within each zone, we model the hypothetical concentration *N* of airborne infectious particles that would be produced by infectious individuals in the space. Consistent with the infection probability distribution suggested by the Wells-Riley equation [37], we model this concentration as “quanta” per volume (unit q/m^3^) where one “quantum” (unit q) of infectious particles is defined as the average amount of infectious particles that will lead to infection in a susceptible person. For brevity, we refer to the concentration of airborne infectious particles as the “quanta concentration”. From this concentration, we calculate the hypothetical exposure rate for susceptible individuals which then produces an estimate of the number of transmission events during the evaluation period.

With this general approach, the main dynamic model for the airborne infectious quanta concentration *C* is as follows:

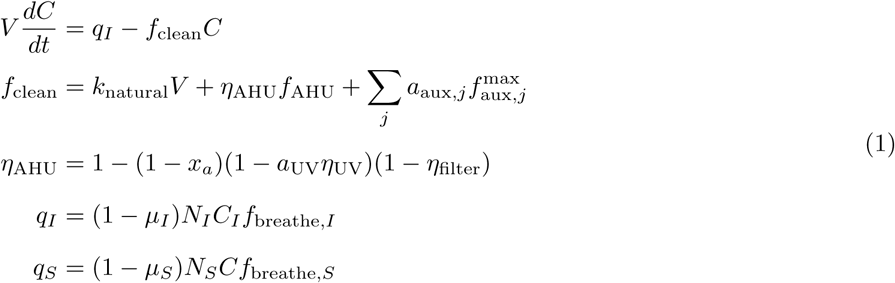

From these variables, we then calculate the primary metrics via

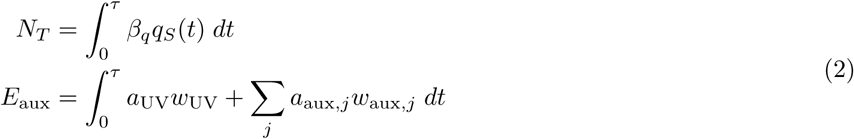

where *N*_*T*_ is the expected number of transmission events that occur and *E*_aux_ the energy consumption associated with UV and auxiliary disinfection, both over the time period *t* ∈ [0, *τ*]. Descriptions and units of all variables are given in Table 1.

**Table 1:**
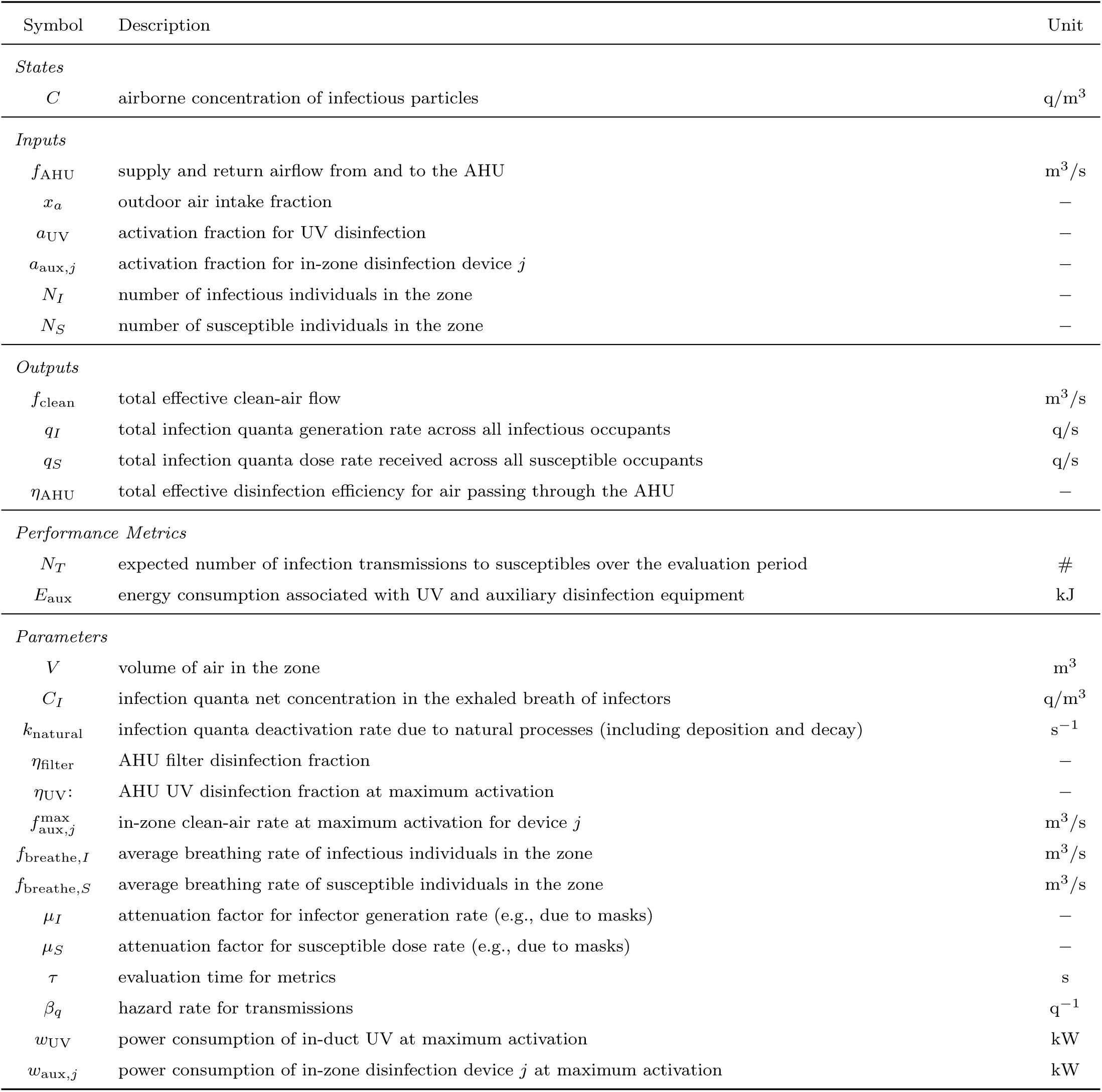
Variables in the infectious particle concentration model.

Although similar to other dynamic formulations of the Wells-Riley equation, the main contribution of this model is that it represents *all* infectious-particle removal sources as an effective clean-air flow *f*_clean_. This value gives the volumetric flow of clean (i.e., *C* = 0) air that would displace infectious particles at the same total rate as the mechanisms being considered. (As mentioned before, similar concepts have been referred to as “clean air delivery rate” [46] and “equivalent outdoor air” [44], but those terms generally refer only to a subset of of the various removal mechanisms.) The key benefits of this quantity are that its definition as a volumetric flow makes it more intuitive for HVAC engineers, and that (in the cases considered) its value is independent of the underlying infectious particle concentration *C*, allowing it to be calculated from readily available building data. In addition, by linearizing the standard exponential infection probability for each individual, we can thus aggregate across occupants and estimate the expected number of transmissions specifically within the specific space being considered. This *space-centric* formulation is critical to guide decision-making, as it allows identification of high-risk spaces and separate optimization of disinfection strategies tailored to each space. Specific details about the model are discussed in Appendix A, including the clean-air delivery calculation, the number-of-transmissions equation, and values for key parameters and inputs.

### 2.2. Thermal Dynamics and Energy Consumption

To model the evolution of temperature and humidity within the zones, we use straightforward (thermal) energy and (water vapor) mass balances respectively. Although there are some pressure fluctuations in the space, they are generally small enough to allow a constant-density assumption for air within the zones. Consistent with the previous infection model, the volume within a zone is assumed to be well-mixed and thus have uniform temperature and humidity. Note that throughout this discussion, we use “humidity” to mean the “absolute humidity ratio” (commonly denoted *ω*) unless otherwise specified.

ODE equations for the mass and energy balances are as follows:

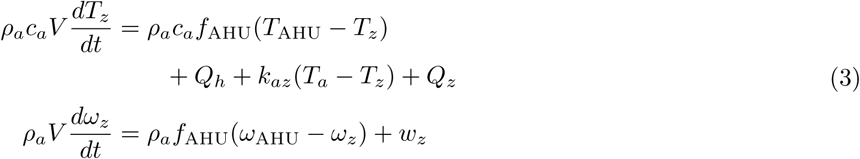

The AHU outlet conditions and loads are calculated from simple heating and cooling coil models with the following functional form (and described in more detail in Section B.2):

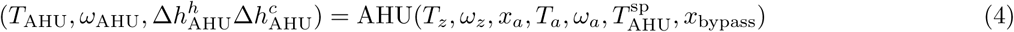

The electricity consumption of the fan is given by a cubic polynomial (see Section B.3 for more details):

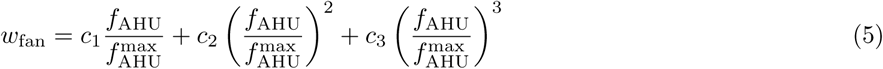

Finally, from all of these outputs we the main metrics associated with energy consumption and thermal comfort in the space:

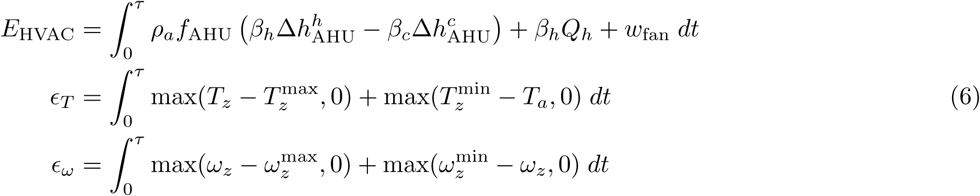

Descriptions and units of all variables in these equations can be found in Table 2.

**Table 2:** Variables in the thermal dynamics and energy model.

| Symbol | Description | Unit |
| --- | --- | --- |
| *States* |  |  |
| $T_z$ | temperature of the air inside the zone | $^{\circ}\text{C}$ |
| $\omega_z$ | humidity ratio of the air inside the zone | — |
| *Inputs* |  |  |
| $f_{\text{AHU}}$ | supply and return airflow from and to the AHU | $\text{m}^3/\text{s}$ |
| $x_a$ | ambient air intake fraction | — |
| $Q_h$ | sensible heating from HVAC heating sources | $\text{kW}$ |
| $T_{\text{AHU}}^{\text{sp}}$ | AHU supply temperature setpoint | $^{\circ}\text{C}$ |
| $T_a$ | ambient temperature | $^{\circ}\text{C}$ |
| $\omega_a$ | ambient humidity ratio | — |
| $Q_z$ | sensible thermal load on the zone | $\text{kW}$ |
| $w_z$ | latent moisture load on the zone | $\text{kg}/\text{s}$ |
| $T_z^{\text{max}}$ | maximum comfortable zone temperature | $^{\circ}\text{C}$ |
| $T_z^{\text{min}}$ | minimum comfortable zone temperature | $^{\circ}\text{C}$ |
| $\omega_z^{\text{max}}$ | maximum comfortable zone humidity | — |
| $\omega_z^{\text{min}}$ | minimum comfortable zone humidity | — |
| *Outputs* |  |  |
| $T_{\text{AHU}}$ | actual supply temperature of AHU | $^{\circ}\text{C}$ |
| $\omega_{\text{AHU}}$ | actual supply humidity of the AHU | — |
| $\Delta h_{\text{AHU}}^h$ | enthalpy difference across the AHU heating coil | $\text{kJ}/\text{kg}$ |
| $\Delta h_{\text{AHU}}^c$ | enthalpy difference across the AHU cooling coil | $\text{kJ}/\text{kg}$ |
| $w_{\text{fan}}$ | electricity consumption at the fan | $\text{kW}$ |
| *Performance Metrics* |  |  |
| $E_{\text{HVAC}}$ | total HVAC energy consumption | $\text{kJ}$ |
| $\epsilon_T$ | total temperature constraint violation | $^{\circ}\text{C s}$ |
| $\epsilon_{\omega}$ | total humidity constraint violation | $\text{s}$ |
| *Parameters* |  |  |
| $\rho_a$ | density of air | $\text{kg}/\text{m}^3$ |
| $V$ | volume of air in the zone | $\text{m}^3$ |
| $c_a$ | heat capacity of air (at average humidity) | $\text{kJ}/\text{kg } ^{\circ}\text{C}$ |
| $k_{az}$ | zone/ambient heat transfer coefficient | $\text{kW}/^{\circ}\text{C}$ |
| $x_{\text{bypass}}$ | coil model bypass airflow fraction | — |
| $c_i$ | fan power curve coefficients | $\text{kW}$ |
| $f_{\text{AHU}}^{\text{max}}$ | maximum flow in the AHU | $\text{m}^3/\text{s}$ |
| $\beta_c$ | energy conversion factor for cooling | — |
| $\beta_h$ | energy conversion factor for heating | — |

For the most part, this model follows standard thermal modeling principles for buildings, with some simplifications to avoid parameters and inputs that are not readily available in real buildings (e.g., accurate solar intensity forecasts or predictions of thermal mass dynamics). We consider both temperature and humidity to ensure that energy consumption is accurately calculated and that occupant comfort is thoroughly considered. Key features of this model are discussed in more detail in Appendix B.

### 2.3. Control Layer

An important feature of the modeling framework is that we consider the actions of the regulatory control layer in response to supplied controller setpoints and the thermal evolution of the zone. This functionality is critical because two of the inputs to the infection model (airflow *f*_AHU_ and outdoor-air fraction *x*_*a*_) that have a significant impact on clean-air delivery *cannot* be directly specified in real buildings and instead are determined as a side effect of the regulatory control layer’s actions. Although the controllers in each piece of equipment operate according to explicitly defined control laws, those laws often consist of complicated rule-based logic that operates on much faster timescales than desired simulation timestep. Thus, rather than try to replicate the control laws directly, we instead choose a phenomenological approach in which we attempt to model the *outcomes* of the controllers’ actions. For example, rather than trying to model the (heavily modified) PI control law within the VAV controllers to specify airflow as a function of setpoint tracking error, we instead assume that the controller is well-designed and thus maintains temperature setpoints within the space whenever possible. This overall approach is similar to the simulation strategy used by EnergyPlus [49], and it simplifies our modeling effort by allowing us to use information about internal system states and disturbances that would not be directly available to the controllers.

The main controller output variables are calculated using the following equations:

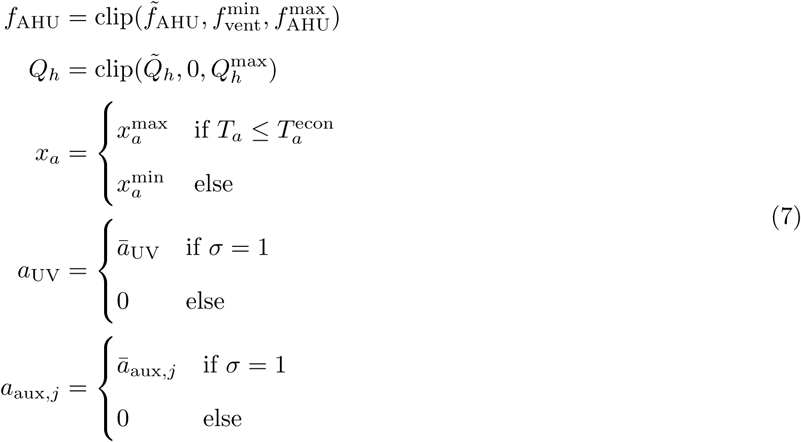

These calculations use the following intermediate variables:

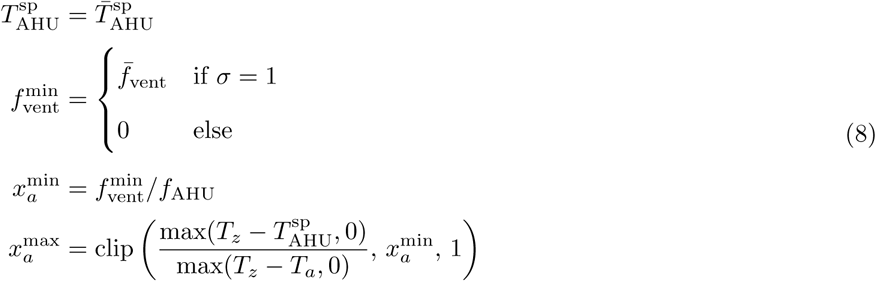

Note that the clip function is defined as clip(*x, a, b*) = min(max(*x, a*), *b*) for *a* ≤ *b*. Variables used in this model are described in Table 3. Specific equations are discussed in the following sections.

**Table 3:** Variables in the HVAC controller model.

| Symbol | Description | Unit |
| --- | --- | --- |
| *Inputs* |  |  |
| $T_z$ | temperature of the air inside the zone | °C |
| $\omega_z$ | humidity ratio of the air inside the zone | — |
| $T_a$ | ambient temperature | °C |
| $\omega_a$ | ambient humidity ratio | — |
| $Q_z$ | sensible thermal load on the zone | kW |
| $T_z^{\max}$ | maximum comfortable zone temperature | °C |
| $T_z^{\min}$ | minimum comfortable zone temperature | °C |
| $f_{\text{AHU}}^{\max}$ | maximum AHU air flow | m<sup>3</sup>/s |
| $Q_h^{\max}$ | maximum reheat rate | kW |
| $\sigma$ | occupancy indicator variable | {0, 1} |
| *Outputs* |  |  |
| $f_{\text{AHU}}$ | supply and return airflow from and to the AHU | m<sup>3</sup>/s |
| $x_a$ | outdoor air intake fraction | — |
| $Q_h$ | sensible heating from HVAC heating sources | kW |
| $T_{\text{AHU}}^{\text{sp}}$ | AHU supply temperature setpoint | °C |
| $f_{\text{vent}}^{\min}$ | minimum outdoor-air ventilation rate | m<sup>3</sup>/s |
| $a_{\text{UV}}$ | activation fraction for in-duct UV disinfection | — |
| $a_{\text{aux},j}$ | activation fraction for in-zone disinfection device $j$ | — |
| $\tilde{f}_{\text{AHU}}$ | hypothetical value of $f_{\text{AHU}}$ that drives $T_z$ to $T_z^{\max}$ in one timestep (see Section C.1) | m<sup>3</sup>/s |
| $\tilde{Q}_h$ | hypothetical value of $Q_h$ that drives $T_z$ to $T_z^{\min}$ in one timestep (see Section C.1) | kW |
| $x_a^{\min}$ | minimum outdoor-air fraction needed to provide minimum ventilation | — |
| $x_a^{\max}$ | maximum outdoor-air fraction that can meet supply temperature setpoint | — |
| *Parameters* |  |  |
| $\bar{T}_{\text{AHU}}^{\text{sp}}$ | default AHU supply temperature setpoint | °C |
| $T_a^{\text{econ}}$ | economizer threshold ambient temperature | °C |
| $\bar{f}_{\text{vent}}$ | occupied outdoor-air minimum ventilation rate | m<sup>3</sup>/s |
| $\bar{a}_{\text{UV}}$ | maximum activation fraction for UV disinfection | — |
| $\bar{a}_{\text{aux},j}$ | maximum activation fraction for in-zone disinfection device $j$ | — |

Overall, the main functionality of the controller model is that it allows prediction of the HVAC control variables that ultimately determine infection risk and energy consumption in the space. In an ideal system, we would be able to manipulate (and thus optimize) the airflow *f*_AHU_ and outdoor-air fraction *x*_*a*_ directly, but in existing HVAC systems, these quantities reflect the response of the regulatory control system and thus can be adjusted only indirectly. Using this simple controller model, we thus bridge the gap between the controller settings we can manipulate and the HVAC variables required for infection-risk and energy modeling. Specifically, by coupling to the thermal model from Section 2.2, we can adjust the static controller parameters *θ* and predict how the HVAC control system will respond. The resulting metrics can then be used to optimize for energy consumption or infection risk depending on our current priorities. Specific controller model components are discussed in more detail in Appendix C.

## 3. Monitoring and Optimization

The models presented in Section 2 have two primary use cases. First, where actual measurements (air flows and temperatures) are available, the models can be used in a monitoring capacity to estimate the clean-air delivery and resulting infection risk within a space. These calculations can be used to determine whether each space is or is not being provided adequate disinfection as actually operated. Second, for spaces with high infection risk or energy consumption, the models can be used for optimization to predict the effects of changing various design or operational parameters. By evaluating the tradeoff between infection risk and energy consumption in a space, decision-making can be guided in accordance with current priorities. We discuss these two use cases in the following sections.

### 3.1. Monitoring

When used for monitoring purposes, we assume that measurements of temperatures and airflows are available from the building management system (BMS). Thus, we need not use the thermal and controller models from Sections 2.2 and 2.3, as the relevant output quantities are already known. We do, however, need to simulate the airborne infection model from Section 2.1 to assess infection risk in the space. The two primary outputs of interest are the expected number of transmissions *N*_*T*_ and the total clean-air delivery rate *f*_clean_; the former can be used to quantify the infection risk in the zone, while the latter indicates to what extent HVAC measures are reducing that risk. Providing both values is important to be able to assess whether HVAC measures are providing sufficient mitigation (as indicated by high *f*_clean_ *and* low *N*_*T*_) or whether non-HVAC operational changes are needed (as indicated by high *N*_*T*_ *despite* high *f*_clean_).

To simulate the infection model, we require values for the airflow inputs *f*_AHU_ and *x*_*a*_, as well as the activation fractions *a*_UV_ and *a*_aux,*j*_ for the relevant disinfection devices. The numbers of infectors and susceptibles *N*_*S*_ and *N*_*I*_ can be chosen from a typical occupancy profile as discussed in Section A.4. If the evaluation period begins during unoccupied hours, then the initial condition for the infection quanta concentration *C* can be set to zero; otherwise, a “warmup” period of roughly 5 hours should be simulated to obtain a more accurate initial condition. From there, the hypothetical infectious particle concentration in the space can be simulated via Eq. (1), and the expected number of transmissions can be calculated using Eq. (2).

Where estimates of energy consumption are required, the AHU and fan models from Eqs. (4) and (5) can be used, since their inputs are generally all known. In addition to *f*_AHU_ and *x*_*a*_, these models also require the zone conditions *T*_*z*_ and *ω*_*z*_, the ambient conditions *T*_*a*_ and *ω*_*a*_, and the AHU conditions *T*_AHU_ and *ω*_AHU_ (although *ω*_AHU_ can also be estimated from the AHU model Eq. (4), as its value is often not measured in real buildings). In-zone heating energy can also be included where *Q*_*h*_ is known, although its value generally cannot be directly measured and must be estimated from other sources (e.g., heating-coil activation commands). We discuss some additional use cases in the following sections.

#### 3.1.1. Uncertainty Quantification

As mentioned in Section 2.1, many of the key infection-model parameters are related to occupant behavior or disease characteristics, which means they are somewhat uncertain in value. For the monitoring procedure described above, we assume mean values of all parameters to simulate mean values of the key infection metrics. However, it may be useful to quantify the uncertainty associated with the model’s predicted number of infection transmissions *N*_*T*_.

Fortunately, for many of the key model parameters, uncertainty quantification is straightforward. First, we note that the infection quanta concentration ODE in Eq. (1) is *linear* in the product of parameters (1 − *µ*_*I*_)*f*_breathe,*I*_*C*_*I*_. Thus, the (time-varying) value of the infection quanta concentration *C* is an *affine* function of this group. We know also that if *C*_*I*_ = 0, then *C* ≡ 0, and thus we have that *C* is *directly proportional* to these parameters. Second, the performance metric *N*_*T*_ is proportional to the product of parameters (1 − *µ*_*S*_)*f*_breathe,*S*_ that multiply *C* to determine *q*_*S*_, and thus its value is *directly proportional* to them. Combining these two facts, we find the relationship

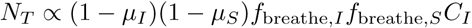

Thus, in order to obtain distributional information for *N*_*T*_, we need only run a single simulation of Eq. (1) to find a baseline value of *N*_*T*_ and then sample its distribution by rescaling the baseline value using samples of the other five parameters.

Unfortunately, the remaining model inputs and parameters are either time-varying or enter the dynamic model nonlinearly. Thus, estimating their impact on the distribution of *N*_*T*_ requires, e.g., Monte Carlo methods that would repeatedly simulate the system. For example, if the number of susceptible occupants *N*_*I*_ has uncertain intra-day variation, then separate simulations should be run for various samples of the *N*_*I*_ *sequence*. Given space limitations, we do not go into more detail here, but simply note that the variance of model predictions can be estimated from the parameter distributions at the cost of running additional simulations.

#### 3.1.2. Disturbance Inferencing

Although the thermal model Eq. (3) is not directly needed for monitoring purposes, it can nevertheless provide valuable insight into the operation of the space. Specifically, we note that the model includes inputs *Q*_*z*_ and *w*_*z*_ for the internal sensible and moisture loads in the space. Although reasonable estimates of these values can be obtained from occupancy profiles and typical energy densities (e.g., from ASHRAE standards [55]), the character of these disturbances can vary significantly from space to space. Thus, to better predict the airflow delivered to each space, it is desirable to have a more accurate forecast of *Q*_*z*_ and *w*_*z*_, which ultimately determine the response of the HVAC system.

To infer the values of these parameters, we note that all other terms in Eq. (3) are generally known with higher accuracy, either via direct measurements from the building or based on simple physical characteristics of the space. In particular, the derivative terms *dT*_*z*_*/dt* and *dω*_*z*_*/dt* can be estimated from data through appropriate filtering. Thus, we can back-calculate values on a pointwise basis as

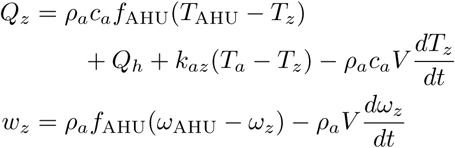

generally with additional filtering to minimize the impact of measurement noise. Once these disturbance trajectories have been calculated, future values can be predicted through various regression techniques [61]. These predicted inputs can then be used for optimization purposes as discussed in the next section.

### 3.2. Optimization

When used for optimization purposes, the dynamic models in Section 2 serve to predict values of key performance metrics that would result from certain decision variables. By coupling the HVAC controller model from Section 2.3 to the thermal model in Section 2.2, we can predict the airflow and energy HVAC energy consumption for a space. The predicted airflow values can then be used as inputs to the infection-risk model from Section 2.1 to estimate infection risk. The key realization is that there is an inherent tradeoff between energy consumption and infection risk in the space: almost all HVAC actions that reduce infection risk (by providing additional clean air to the space) also result in increased energy consumption. Since there are multiple different mechanisms by which clean air can be provided, the goal of the optimization is to identify the most efficient clean-air sources, which can vary from day to day based on ambient conditions. In addition, it may be the case that some spaces already provide adequate disinfection under standard operation, and thus there is no motivation to further increase clean-air delivery.

In order to address these needs, we formulate a multiobjective optimization problem using the dynamic models to predict the necessary decision metrics. Mathematically, the overall structure of the optimization problem is as follows:

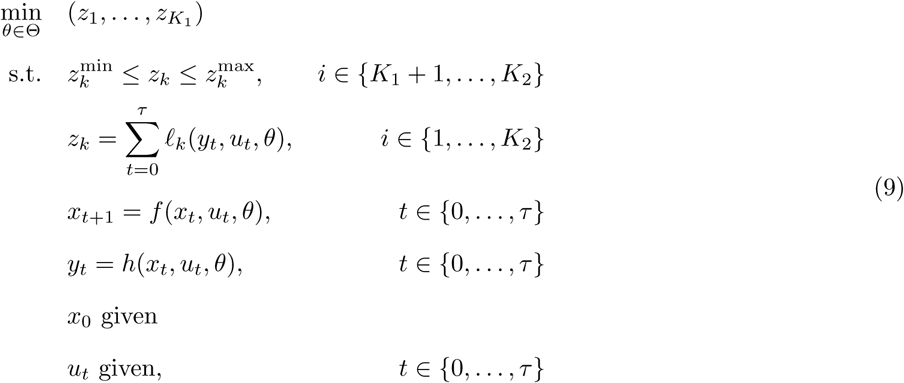

That is, we perform multiobjective optimization over a set of model parameters *θ* ∈ Θ with each scalar objective function and constraint *z*_*k*_, calculated from the inputs *u*_*t*_ and outputs *y*_*t*_ of the simulation model. Note that in this context, the models *f* (·), *h* (·), and *ℓ*_*k*_ (·) represent the *coupled* models, i.e., the combination of infections quanta, thermal, and controller models as mentioned in the previous paragraph.

Owing to the multiple objectives, the goal is not to find just a single optional point *θ*^∗^ but rather the *set* of non-dominated points

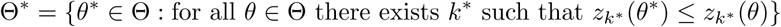

also referred to as the “Pareto set”. All values in this set are efficient in the sense that it is not possible to improve the value of one objective function without making a different objective function worse. The specific values to implement can then be chosen from this set in accordance with current infection-risk and energy priorities.

The proposed optimization process is illustrated in Fig. 1. As will be discussed in Section 3.2.3, we choose a *finite* set of optimization cases specified by the values of their decision variables. A simulation is run for each optimization case, holding the other model parameters and inputs constant across all cases. Within the simulation, the controller and thermal models (from Section 2.3 and Section 2.2 respectively) are coupled, with the controller model determining the HVAC control actions (primarily flow rate and outdoor air fraction) based on the previous zone state (i.e., temperature and relevant disturbances) and configured setpoints. The thermal model then evolves based on those control actions, and the process is repeated until the end of the simulation period. In parallel, the HVAC control actions are passed to the infection model so that infection risk and other objectives can be evaluated. Once the objectives and constraints have been simulated for each optimization case, a Pareto search is performed to characterize the feasibility and optimality of each case. The optimal cases can then be presented the user to select from in accordance with their current operating goals. We discuss the decision variables, performance metrics, and Pareto optimization algorithm in the following sections.

**Figure 1:**
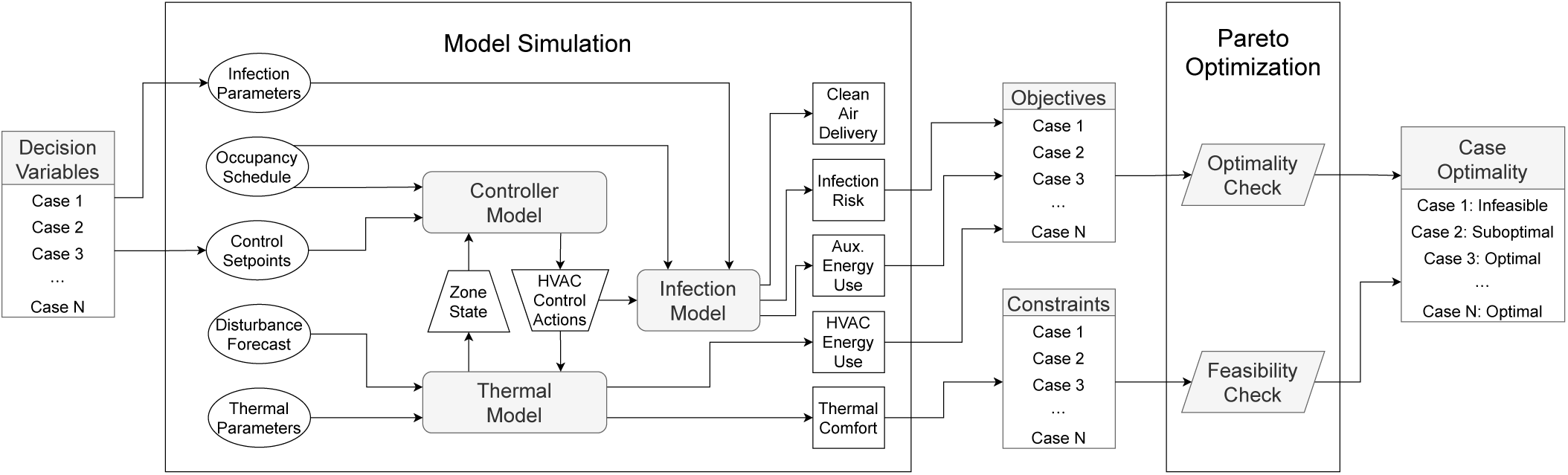
Data flow diagram for optimization process. For each optimization case, the corresponding decision variables are simulated using the proposed models to calculate values of objective functions and constraints. Results are then post-processed via the Pareto Optimization algorithm to identify whether each case is infeasible, suboptimal, or optimal.

#### 3.2.1. Decision Variables

For the purposes of optimization, we split the decision variables into two different groups: operational variables that can be adjusted via setpoints sent to the BMS, and design variables that would require physical equipment to be installed or replaced. The key distinction is that operational variables can be reasonably be adjusted on a daily or faster basis, whereas changes to design variables would generally be changed only monthly or less frequently. Thus, the frequency of optimization (and the time period considered in the optimization) will typically depend on the variables being adjusted. Fig. 1 illustrates the general case in which the decision variables can affect both the HVAC control setpoints and the parameters of the infection model, but often only one group (i.e., operational or design) of variables will be optimized at a time.

The primary operational variables to optimize are the supply temperature setpoint 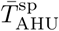, the minimum ventilation setpoint 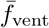, economizer threshold temperature 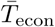, and the activations *ā*_UV_ and *ā*_aux,*j*_ for disinfection devices. Note that all of these variables are parameters of the controller model from Section 2.3. For simplicity, we assume that the operational variables are all held constant over the simulation interval (e.g., each day) so as to keep the optimization space reasonably small. Various extensions are possible that would adjust these variables more frequently (e.g., every hour), but care would be needed to account for the transient dynamics of the control layer after setpoint changes, as these effects are not included in the current models.

The primary design variables to optimize are the in-duct filter type, which implicitly adjusts *η*_filter_ and the fan-power coefficients *c*_*i*_, and the presence of in-zone disinfection devices, which would add elements *j* included in the clean-air delivery and disinfection energy calculations of Eqs. (1) and (2). Of course, which values are included in the optimization space will need to account for the physical constraints of the HVAC system (e.g., certain filter types may not be compatible with the filter rack present in the duct).

#### 3.2.2. Objective Function and Constraints

Once the set of decision variables has been chosen, the next step is to choose the objective functions and constraints to include in the optimization problem. For our purposes, we are interested in multiobjective optimization to evaluate the tradeoff between energy consumption and infection risk in the space. The corresponding performance metrics are the expected number of transmissions *N*_*T*_ (from Eq. (2)) and the total energy consumption *E* = *E*_HVAC_ + *E*_aux_ (from Eqs. (2) and (6)). The goal of the optimization is not only to minimize these metrics separately, but also to identify the Pareto set in the two-dimensional objective space to identify the efficient set of decision variables.

Although the decision space is already bounded based on the chosen optimization ranges for the optimization variables, it is also important to ensure that selected values do not have unintended consequences for the space. For example, although a given AHU may physically be able to supply air at a temperature 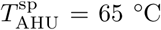, this value may not provide adequate dehumidification of the supply air, which could thus lead to unacceptably high humidity within the zone. To remove such variables from the decision space, we thus include the thermal and moisture comfort metrics *ϵ*_*T*_ and *ϵ*_*ω*_ from Eq. (6) as explicit constraints in the optimization problem. Specifically, thresholds are set on both metrics, and if a particular set of variables leads to either threshold being violated, then that set of variables is deemed infeasible and not part of the Pareto set. Note that in extreme circumstances (e.g., if the comfort bounds 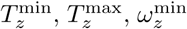, and 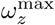 have been chosen too tightly), then there may not be any feasible points left in the optimization space. Thus, to obtain a solution in such cases, it may be necessary to iteratively relax the constraint-violation thresholds until at least one feasible point is identified. However, for well-designed systems, such infeasibility is not expected to be an issue.

Both objective functions and constraints are calculated from the outputs of the simulation models as illustrated in Fig. 1. Each of these metrics aggregates over the simulation interval to produce a single scalar value from the time-varying model outputs. These values are then processed via the Pareto optimization algorithm as described next.

#### 3.2.3. Pareto Optimization Algorithm

Although there is a wealth of literature on multiobjective optimization (and optimization in general), we propose in the interest of usability a fairly simple-minded optimization algorithm for our problem. Specifically, we choose a *finite* set of points 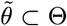 that approximates the full space and then identify the Pareto set 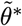 from just that finite set by simulating the performance metrics for *each* point within that set. This choice is appropriate for the following reasons:

- The number of independent decision variables is generally between 3 and 5, which means a simple grid over the continuous variables will not give a prohibitive number of points.
- Some decision variables (e.g., filter type or on/off choice for equipment) are implicitly discrete, and thus continuous optimization methods cannot be applied in a straightforward manner.
- Enumerating the full set of points avoids the need to calculate gradient information, as optimization can be performed simply by looking at the simulated metrics for each of the finite set of points.

For other problems where these properties do not hold, it may be necessary to use more sophisticated optimization algorithms for the problem to remain tractable. However, as we will show in the examples, this strategy is suitable for the current problem.

To build the set 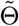, we simply choose a finite set of cases for each scalar decision variable and take 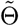 to be their Cartesian product. For variables that are discrete, the finite set is already defined, while for continuous variables, we choose a minimum and maximum value along with a handful of values in between. For example, suppose our decision variables are the supply temperature setpoint 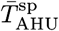, the minimum ventilation rate 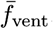, and whether to activate UV disinfection *ā*_UV_. We would thus define the set

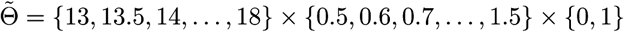

i.e., choosing 11 values between 13 °C and 18 °C for 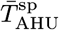, 11 values between 0.5 m^3^/s and 1.5 m^3^/s for 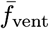, and two values for *ā*_UV_ corresponding to “on” and “off” respectively. Thus, to optimize over this set, we need perform 11 × 11 × 2 = 242 simulations.

Once the set 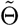 has been chosen, the overall optimization algorithm is straightforward:

1. Initialize the Pareto-optimal set 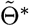, the suboptimal set, 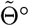, and the infeasible set 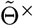 to the empty set ∅.
2. For each 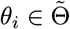:
  2.1. Simulate the model using parameters *θ*_*i*_ to determine each *z*_*k*_(*θ*_*i*_).
  2.2. If all bounds 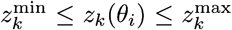 are satisfied for *k* ∈ {*K*_1_ + 1, …, *K*_2_}, add *θ*_*i*_ to 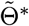. Otherwise, add *θ*_*i*_ to 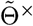.
3. For each 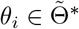:
  3.1. For each 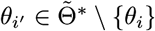:
    3.1.1. If all *z*_*k*_(*θ*_*i′*_) ≤ *z*_*k*_(*θ*_*i*_) for *k* ∈ {1, …, *K*_1_}, remove *θ*_*i*_ from 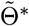 and add to 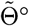.
4. Return the Pareto-optimal set 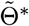, the suboptimal set 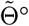, and the infeasible set 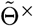.

Thus, we require one model simulation for each case in 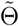 and *K*_1_ comparisons for each *pair* of cases in 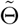. Note that more efficient algorithms are known [62], but this naive implementation is sufficient for the problem sizes we consider. In terms of Fig. 1, Step 2.1. is the model simulation, Step 2.2. is the feasibility check, and Step 3.1.1. is the feasibility check. The returned sets 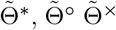 (representing the Pareto-optimal, suboptimal, and infeasible subsets) are thus disjoint and characterize each case in in the original set 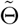.

Once the approximate Pareto-optimal set 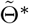 has been determined, the only decision left is to choose a specific point to implement within the system. This selection could be made via thresholds on the variables, e.g., choosing the solution with the lowest energy consumption *E* such that the number of transmissions *N*_*T*_ ≤ 1 if any such points exist, or otherwise choosing the solution with the lowest *N*_*T*_. The exact selection criteria will depend on a building manager’s current priorities, but by providing the full Pareto set, the fundamental tradeoff can be fully evaluated. Suboptimal and infeasible points can also be explored as desired, but they generally should not be implemented.

## 4. Illustrative Examples

In this section, we show how the proposed models and optimization framework can be used to generate insights and guide operational decisions in buildings. We split the examples into two main categories, the first using data obtained from real buildings (during pre-pandemic operation) and the second using data obtained from building simulations. For both categories, we note that the associated observations and discussion are intended as specific for the exact buildings and spaces being illustrated. In particular, calculated infection rates for a particular example space should not be interpreted as typical values for all spaces of that type in other buildings, as specific characteristics can vary significantly. Indeed, a key point of the examples is that energy and infection metrics depend on many individual factors, and thus the quantitative analysis proposed in this paper is necessary to achieve consistent positive outcomes.

### 4.1. Real Data for a Commercial Building

To illustrate the potential uses of our proposed modeling and optimization framework, we start by applying the models to real building data. For these examples, we will use datasets from two zones in a real building: (1) an “Office” zone consisting of 25,000 ft^2^ of open-plan office floorspace (with some conference rooms and small offices) served by the same AHU; and (2) a “Gym” zone consisting of 5,000 ft^2^ of floorspace containing a weight room, an aerobics studio, and locker rooms. Both datasets come from the same commercial building in the US Midwest, and they cover the time range from August 2019 through March 2020 (after which point the building was closed due to the COVID-19 pandemic). Data was collected at a 1-minute frequency and then re-sampled to 15-minute intervals for our purposes. The Office zone is nominally occupied from 6am to 7pm on weekdays and closed on weekends, while the Gym zone is occupied from 5am to 9pm on weekdays, 7am to 8pm on Saturdays, and 7am to 2pm on Sundays.

As discussed in Section 2.1, calculation of infection risk requires values of key parameters relating to occupant activity. For the Office zone, we assume a breathing rate of 0.7 m^3^/h, while in the Gym zone, we use a higher breathing rate of 1.6 m^3^/h. These values correspond to “low” and “moderate” activity respectively for a typical adult occupant [63]. We note that there is likely to be high variance in the breathing rate for Gym due to the different activities (aerobic exercise, weightlifting, calisthenics, resting, etc.), so our chosen value is best interpreted as a time-averaged mean. For the exhaled quanta concentrations in the zones, it reasonable to suspect that Gym occupants have a higher overall exhaled particle concentration, as they are more likely to be breathing deeply and through their mouth compared to Office occupants. However, it is also plausible that actively infectious individuals would be less likely to exercise if they are feeling symptomatic, and thus we would expect Gym infectors to have below-average viral load (and also exhaled quanta concentration). Given this uncertainty and the absence of definitive data, we use the same mean exhaled quanta concentration of 20 q/m^3^ in both spaces, corresponding to light vocalization [4].

Finally, the models require a time-varying occupancy profile, which is unfortunately not provided by the dataset. Thus, for both zones, we use a surrogate profile obtained by rescaling default schedules from ASHRAE [55] using design occupancy, which is taken as 5 and 10 occupants per 1000 ft^2^ respectively [54]. Zone areas and volumes are taken from floorplan diagrams, and we assume MERV8 filters with no occupants wearing masks, as is consistent with pre-pandemic operation.

#### 4.1.1. Data Trends and Metrics

To begin analysis, we start by examining the daily trends inherent in the datasets. The key BMS measurements used in the infection-risk model are the outdoor air ventilation (which contributes directly to clean-air delivery) and the total supply air flow (which provides clean-air via filtration). To illustrate daily and weekly trends, Fig. 2 shows values of these variables along with the assumed time-varying occupancy profiles. For the Office zone, we see that total supply air flow is reasonably constant throughout the occupied period on a given day, but that the amount of flow provided does change from week to week, likely corresponding to seasonal changes. The provided ventilation varies much more significantly, perhaps due to the action of the economizer, but it still covers a fairly narrow range. By contrast, we see that there is much greater variation in the Gym zone, both within a given day and from week to week. We almost always see a large spike in airflow at the beginning of the occupied period, which indicates that the zone heats up overnight and thus the temperature controllers take aggressive action as soon as they are activated in the morning. There are also some other flow spikes likely associated with extra occupancy around lunchtime and at the end of the workday (but unfortunately the occupancy profile does not reflect those features). We do see that the ventilation varies much more significantly, which indicates the presence of measurement noise in the sensor but could also be caused by operation of the economizer. Consistent with ASHRAE standards, we see higher (normalized) airflows in the Gym zone compared to Office. We note also that in accordance with the operation of the building, there is zero occupancy in the Office zone on the weekends, but the Gym zone remains open. Thus, the HVAC system is inactive in the Office zone but active in the Gym zone during these days.

**Figure 2:**
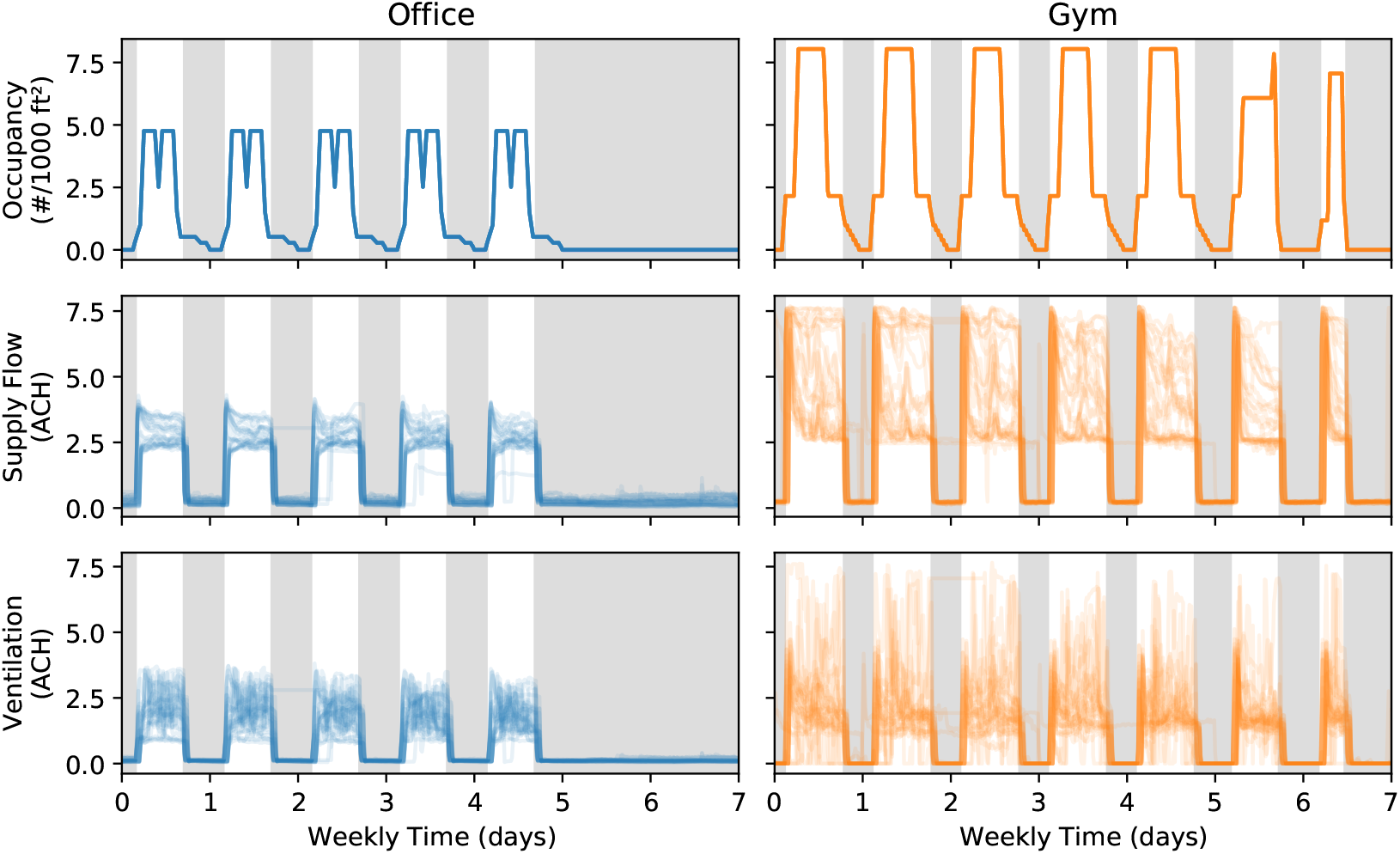
Weekly flow data and occupancy profiles for the real building zones. Plots show 26 overlaid weekly profiles (with time zero corresponding to midnight on Monday). Shaded regions correspond to unoccupied hours when the HVAC system is deactivated.

To assess the infection risk present in these zones, we take the data from the previous plot and simulate the dynamic infectious-particle models from Section 2.1 to calculate average clean-air delivery rate and expected transmissions for each day of operation. Given the uncertainty in the exhaled quanta concentration, we also show a shaded confidence interval corresponding to exhaled quanta concentrations between 5 q/m^3^ and 80 q/m^3^ (with the points assuming the mean value of 20 q/m^3^). These values are plotted in Fig. 3. From these results, we can make three key observations. First, we note that infection risk in both zones is lower on weekends than on weekdays despite similar or significantly lower clean-air delivery rates. This behavior simply mirrors the occupancy cycles of the respective spaces, with lower weekend occupancy for the Gym and zero weekend occupancy for the Office. Second, we note that although the clean-air delivery to the Gym zone is almost always higher than that of the Office zone, the average infection risk for the zone is nearly 10 times higher than that of the Office zone. This discrepancy is explained by the occupancy differences in the two spaces: in the Gym, there is a higher density of people who are all breathing at elevated rates. (Indeed, the Gym has a 2.3 times higher breathing rate and roughly a 1.5 times higher occupant density, which suggests a 2.3^2^ × 1.5 ≈ 7.9 times higher infection rate, all other effects equal.) Thus, although the HVAC system removes more of the particles, there is an inherent higher infection risk compared to the Office zone where occupants are quietly seated at their desks. Third, we note that clean-air delivery rate is generally higher in warmer months and lower in colder months. This behavior is consistent with the fact that supply air flows are elevated in the cooling season to mitigate additional heat loads, whereas in the heating season, flows are lower and may sit at their configured minimum bounds. Under steady-state conditions, transmission rate is inversely proportional to clean-air delivery rate, and thus the observed 40% decrease in clean-air delivery for the Gym zone roughly corresponds to the observed 65% increase in transmissions when comparing the winter months to the summer months.

**Figure 3:**
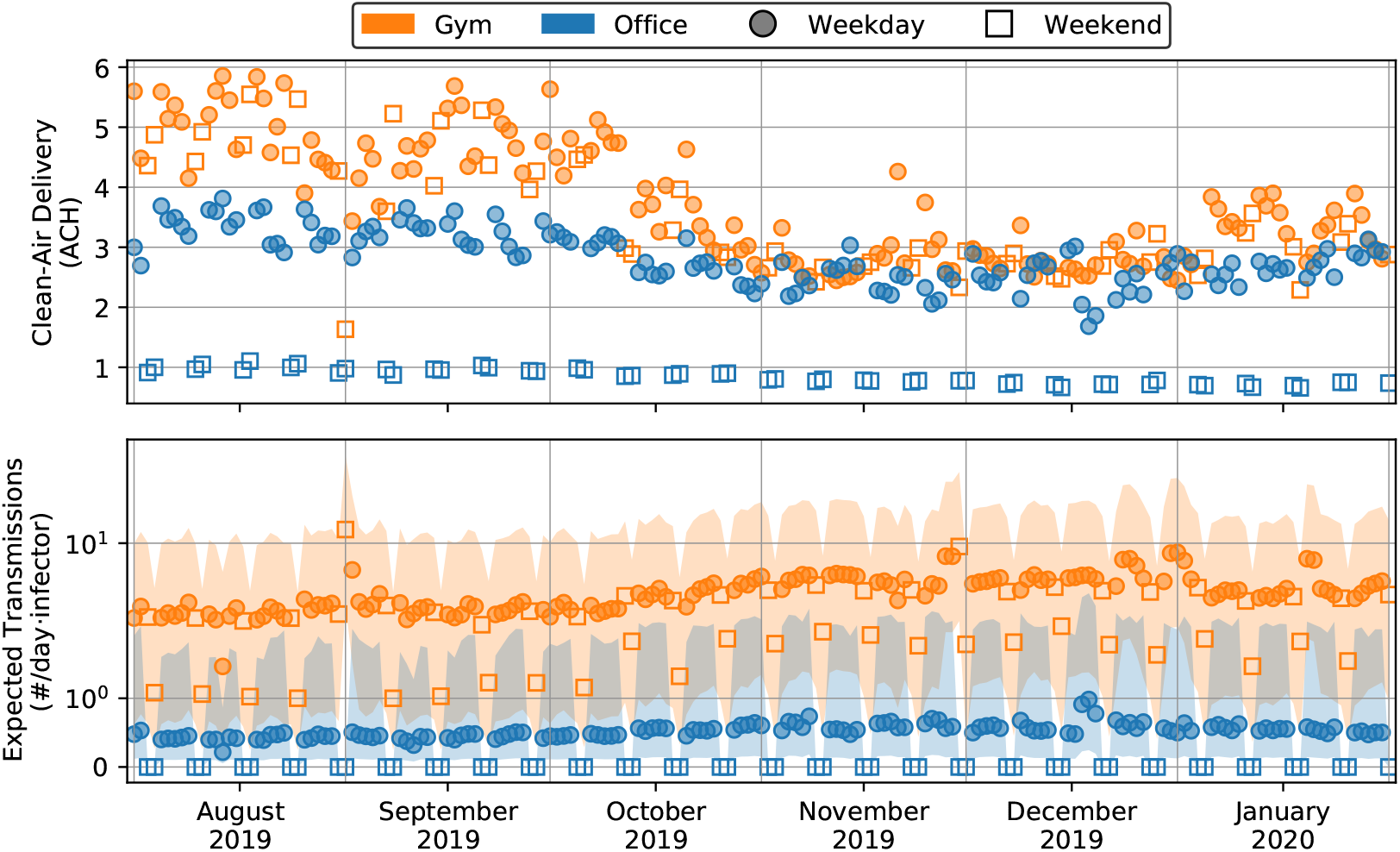
Daily average clean-air delivery and expected transmissions for the real building zones. Each point reflects the average value of the metric over a 24-hour simulation. For transmissions, points are calculated assuming a mean exhaled quanta concentration of 20 q/m^3^, while the shaded region shows the resulting range for exhaled quanta concentrations between 5 q/m^3^ and 80 q/m^3^

Before moving on, we briefly discuss the uncertainty associated with the two variables. For the clean-air delivery rate, the only uncertain terms are the passive decay rate for the infectious particles (due to both deactivation and deposition) and the filtration efficiency for the in-duct filter. Because both effects exhibit particle-size dependence, their actual effect will depend slightly on the specific particle distribution produced by the infector. However, these values are reasonably well characterized, and their range of variation is generally small. The remaining terms in the clean-air calculation are flow measurements taken directly from the BMS. Although flow sensors can be very noisy, we expect only minuscule error when averaged over the course of a full operating day. Thus, we plot the clean-air delivery without an uncertainty region. By contrast, the infection rate depends linearly on the exhaled quanta concentration, which is known to vary by up to three orders of magnitude depending on vocalization level and mode of respiration, from nasal breathing to speaking and singing [4]. Accordingly, we have shown an uncertainty range corresponding to a factor of 4 above or below the chosen mean value. A key takeaway is that, although the baseline infection risk in the Gym is inherently higher than in the Office, this analysis does *not* suggest that Gym zones are always highly dangerous or that Office zones are always a safe haven. Instead, it is necessary to examine both types of spaces on a case-by-case basis to determine the correct course of action.

#### 4.1.2. Prediction and Optimization

We turn now to showing how these models can be used for the purposes of prediction and optimization. In this use case, we wish to predict the operation of the HVAC system one day in advance, which means we do not have access to flow measurements from the BMS and must predict them using the controller and zone-temperature models. Thus, we start by showing example day-ahead predictions of clean-air delivery using the models from Section 2.

Important inputs to the thermal models include the internal heat and moisture disturbances. To predict these values in advance, we use the following procedure: back-calculate disturbances on the prior 7 days of data as discussed in Section 3.1.2, filter that trajectory by retaining only the most dominant Fourier modes in the frequency domain, use the first day of that trajectory as the disturbance for the simulation. This procedure implicitly assumes that successive weeks follow approximately the same disturbance trajectories. Certainly more sophisticated disturbance-prediction algorithms are possible, but even this extremely simple strategy produces adequate results on the test data. In Fig. 4 we show day-ahead clean-air delivery predictions for each zone under summer and winter conditions. Even though these predictions are far from perfect, they nevertheless capture the general daily trends in airflow and thus produce sufficiently accurate infection-risk predictions. As a result, the models are suitable for use in optimization.

**Figure 4:**
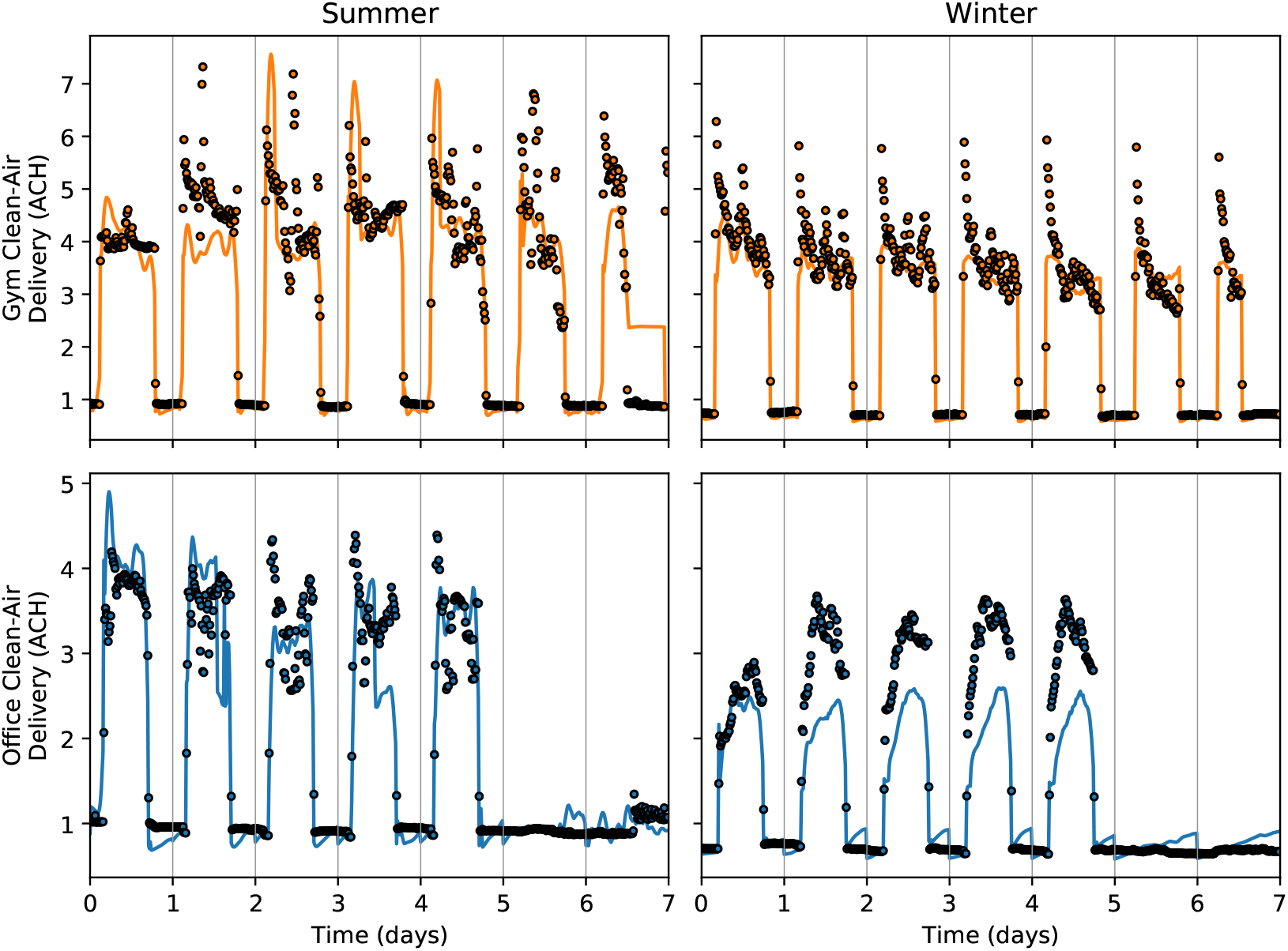
Day-ahead clean-air delivery predictions using the full dynamic models. Points show values computed from the actual BMS data, while lines show values predicted by the model. Data for Summer are taken from August 2019, while data for Winter are taken from February 2020.

To close this example, we present some sample optimization results using the data just presented. In each of the four cases, the optimization period covers 24 hours of operation (corresponding to the the first day from each case in Fig. 4). The decision variables are the supply temperature setpoint (sampled at 25 points between the bounds of 55 °F and 65 °F) and whether or not to activate 2 ACH of supplemental in-zone filtration (provided by standalone filtration devices) during occupied hours. The two primary objective functions are energy cost and expected transmissions, and the constraints are maintenance of zone temperature and humidity within comfortable ranges during occupied hours. Following the optimization strategy presented in Section 3.2, each case thus runs 50 simulations (corresponding to the different values of the decision variables) of the dynamic models from Section 2, and then the Pareto search described in Section 3.2.3 is applied to categorize each solution. These results are shown in Fig. 5. Note that in both cases, the number of transmissions is determined using the mean exhaled quanta concentration of 20 q/m^3^. The effect of uncertainty in this parameter would be to uniformly scale the calculated value up or down, which does not affect the optimality characterization of the points.

**Figure 5:**
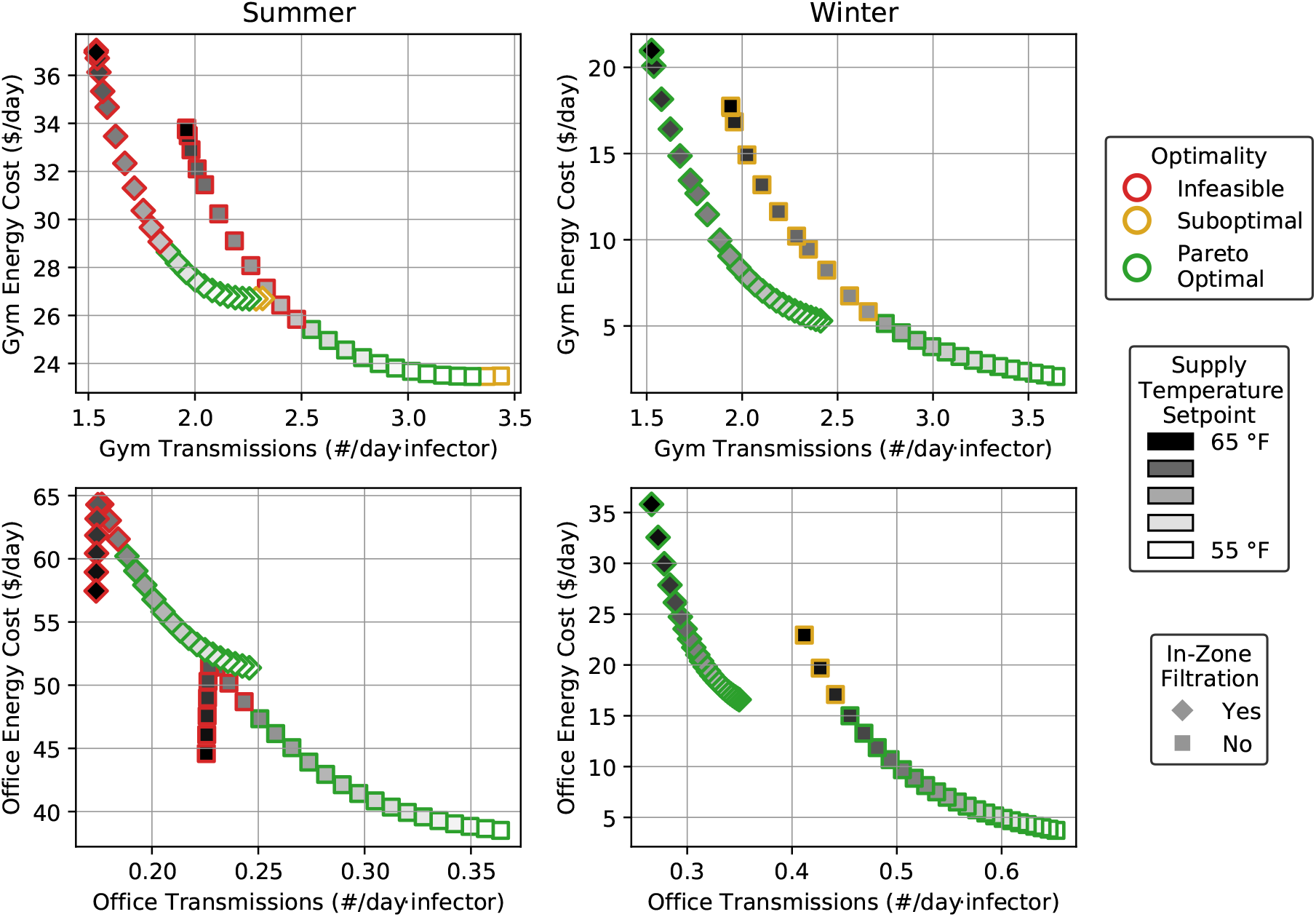
Optimization results for the real building zones under two different weather conditions. Disturbance and weather data for each case are taken from the first day of the corresponding case in Fig. 4.

From the sample optimization results, we see that both decision variables decrease the airborne transmission rate and generally increase energy consumption as their values are increased. For supply temperature setpoint, there are some regions where energy cost is reduced with increasing supply temperature (which corresponds to conditions where the reduced latent load is larger than the increased fan power), but they usually cannot be exploited. A key observation is that in the summer conditions, it is not possible to increase the supply temperature setpoint all the way to the established upper bound, as the reduction in de-humidification at the AHU cooling coil eventually leads to violation of the humidity comfort constraint. By contrast, during winter, the full range of supply temperature setpoints is available, as the outdoor air does not need to be de-humidified. In all cases, when choosing a course of action, only Pareto optimal solutions should be considered, since feasible but suboptimal solutions consume unnecessary energy to achieve a given infection rate (or vice-versa). Thus, as priority shifts from energy to infection risk, the general strategy is to increase supply temperature up to a certain point (which depends on weather conditions) and then activate the supplemental in-zone filtration units (while dropping the supply temperature back to its minimum value) before increasing the supply temperature setpoint once again to achieve maximum clean-air delivery. Overall, we see that even with just these two decision variables being considered, the range of achievable infection rates (and the most energy-efficient strategy to achieve them) depends on both space characteristics and outdoor conditions. Therefore, it is important to update operational decisions regularly to account for changes to these variables.

### 4.2. EnergyPlus Simulations

In our second set of examples, we use simulated building data produced from EnergyPlus [49] using typical meteorological year (TMY) weather data. Although the EnergyPlus simulated data is not as realistic as the real data in the previous section (in particular it is much less noisy and is subject to much more regular disturbance profiles), the key benefit is that we can directly compare the effects of different control actions. Specifically, for a given day’s weather, we can simulate *all* candidate sets operational variables rather than just the default operating policy. We will use this feature to more thoroughly assess the available control space and the accuracy of our models.

For this purpose, we consider the first floor of the “Large Office” reference building provided by EnergyPlus, which consists of 38,000 ft^2^ of floorspace served by the same AHU with a design occupancy of 192 occupants. HVAC equipment in the building is auto-sized by EnergyPlus using the chosen TMY weather data. We assume the same occupant breathing rate of 0.7 m^3^/h and exhaled quanta concentration of 20 q/m^3^ as in the previous example. Similarly, we assume MERV8 in-duct filters and that occupants are not wearing masks. For optimization variables, we allow the minimum ventilation rate to be varied from its default level (chosen by EnergyPlus in accordance with ASHRAE standards) to double that value, while we allow the supply temperature setpoint to be adjusted from its default of 55 °F up to 65 °F.

#### 4.2.1. Metric Trends

To begin the simulation analysis, we examine the values of our two main objective functions and how they vary as the operational variables are changed. For each of the four extreme points in the optimization space, we simulate a full year of the building operating with those variables using EnergyPlus to obtain the required BMS data. We then use that BMS data with the proposed models to calculate daily values for energy cost and infection rate. We perform this process for both a cold climate (TMY data from Chicago, IL) and a hot climate (TMY data from Houston, TX). The results of these simulations are shown in Fig. 6. For the purposes of energy accounting, we assume cooling is provided via electricity at a COP of 3, while heating is provided by gas at 90% efficiency. We use an electricity price of 0.12 $/kW h and a gas price of 0.80 $/therm. For brevity, we do not show thermal discomfort, although all temperature comfort violations are below 4 °F · h/day, and humidity comfort violations are generally confined to the winter with indoor conditions being too dry. To illustrate the relative efficiency of the two operational variables for reducing infection risk, we also plot the cost of extra clean air. This value is calculated as

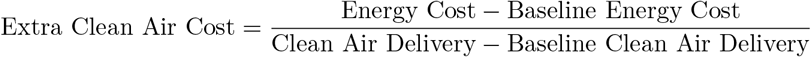

which gives the per-unit cost of the extra clean air provided relative to baseline operation. Lower values of this metric indicate a more efficient source of clean air.

**Figure 6:**
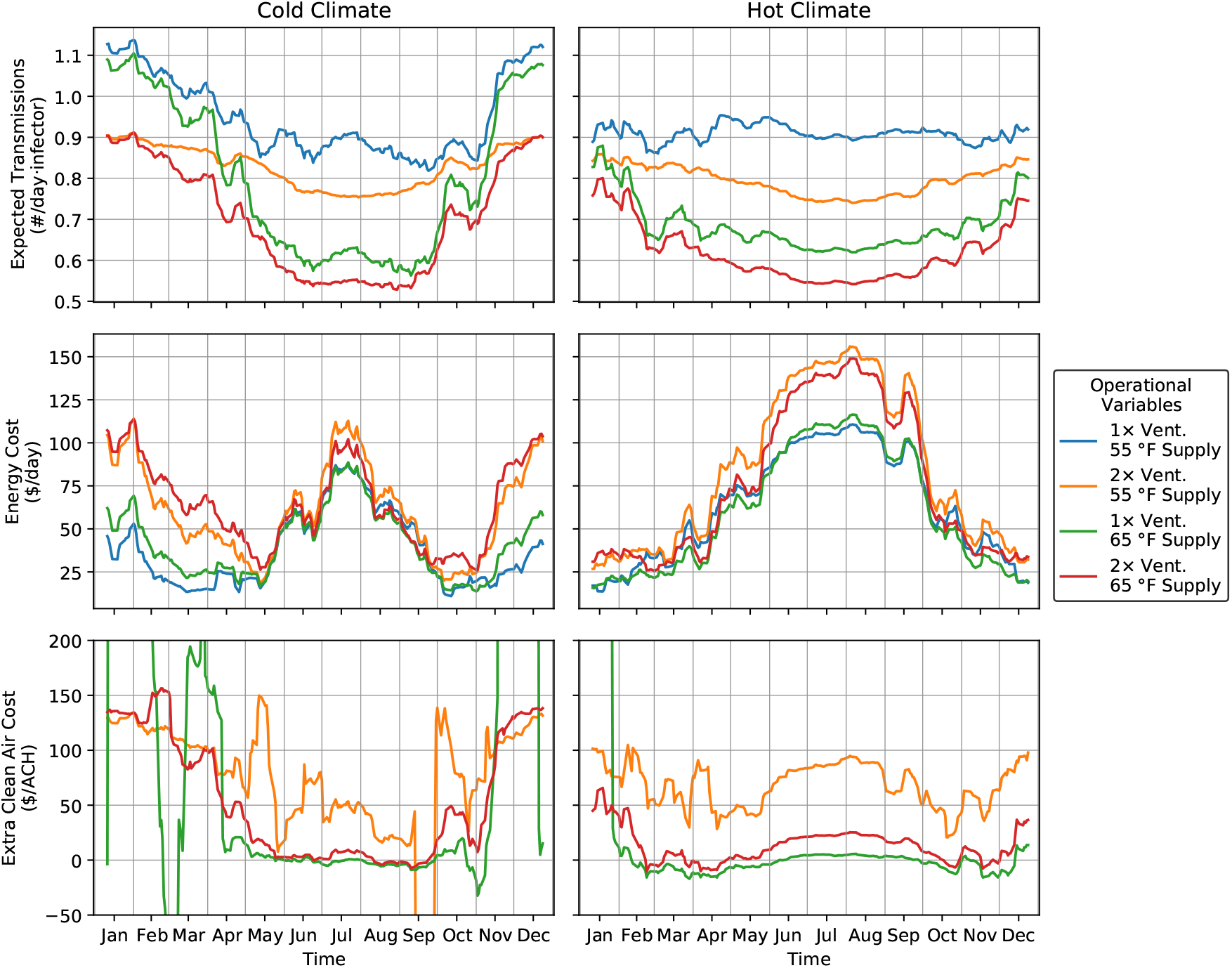
Daily metrics calculated from EnergyPlus simulation data. Lines show ten-day rolling averages for non-holiday weekdays. Values of operational variables are the four extreme points in the optimization set. Extra clean air cost is calculated relative to the baseline operation with both operational variables at their minimum value.

From these results, we see similar behavior as in the previous real-data examples, but with new trends emerging from the new optimization variable. First, we note once again that infection rate is generally higher in the winter months and lower in the summer months, simply due to variation in the average airflow provided by the HVAC system. We note that this effect is much more pronounced in the Cold climate compared to Hot due to the weather-dependent cooling loads (and resulting airflow differences). Second, we see that increased ventilation almost always requires additional energy consumption, and the magnitude is much higher in particularly hot or cold weather. The explanation for this effect is that because the building is equipped with an economizer using an aggressive suitability threshold, the system will automatically increase ventilation above its minimum value when energetically favorable. Thus, any additional ventilation that results from increasing the minimum ventilation setpoint necessarily requires an energy penalty. By contrast, increasing the supply temperature setpoint can potentially *reduce* energy consumption, and so it tends to be much less costly. Finally, we note that the HVAC system by itself has a somewhat narrow range to reduce the infection rate. Comparing the maximum and minimum infection rates on each day, we see less than a factor of two reduction. Thus for spaces with unacceptably high infection rate, it may be necessary to install standalone-filtration or other disinfection devices to achieve infection-risk goals.

For the purpose of decreasing infection risk, we see that increasing the supply temperature setpoint is less effective than increasing the ventilation rate in the heating season but more effective in other seasons. When choosing between the two, we generally prefer the variable with the lower cost of extra clean air. The bottom plot in Fig. 6 shows that the extra clean air provided by increasing the supply temperature setpoint costs next to nothing throughout much of the year, but during the heating season, its cost can jump to more than $200/ACH. This observation is consistent with the fact that the HVAC systems often operate near minimum-flow constraints during the heating season due to the small cooling load, and thus adjusting the supply temperature setpoint has little to no effect on clean-air delivery but a large effect on energy cost (since extra heating is required to meet the higher supply temperature). However, adjusting the supply temperature setpoint in the cooling season can have a significant impact on clean-air delivery, with only a minor change to energy consumption. By contrast, Fig. 6 shows that doubling the ventilaion rate yields a more consistent cost between $50/ACH and $100/ACH for extra clean air throughout the year, which tracks with the changing weather conditions throughout the year. Overall, this analysis illustrates that the two operational variables can have very different effects on energy cost and infection risk throughout the year, which motivates the proposed optimization approach to ensure that the most efficient operational variables are selected based on current weather conditions.

#### 4.2.2. Tradeoff Curves

Given the trends observed in the previous section, we now look at the optimization tradeoffs in more detail. For each of the simulation cases, we choose a representative day for each season and simulate our dynamic models using the appropriate parameters and weather data. Specifically, we grid over the optimization space (supply temperature between 55 °F and 65 °F; minimum ventilation rate between 1 and 2 times the default value), simulate one day of HVAC operation (using the proposed models from Section 2) for each grid point, and plot the resulting energy consumptions and infection rates. These results are shown in Figs. 7 and 8. A key observation is that the shapes of the mapped optimization sets (and thus the Pareto-optimal points within those sets) vary significantly based on the weather, and thus optimal operation changes as well.

**Figure 7:**
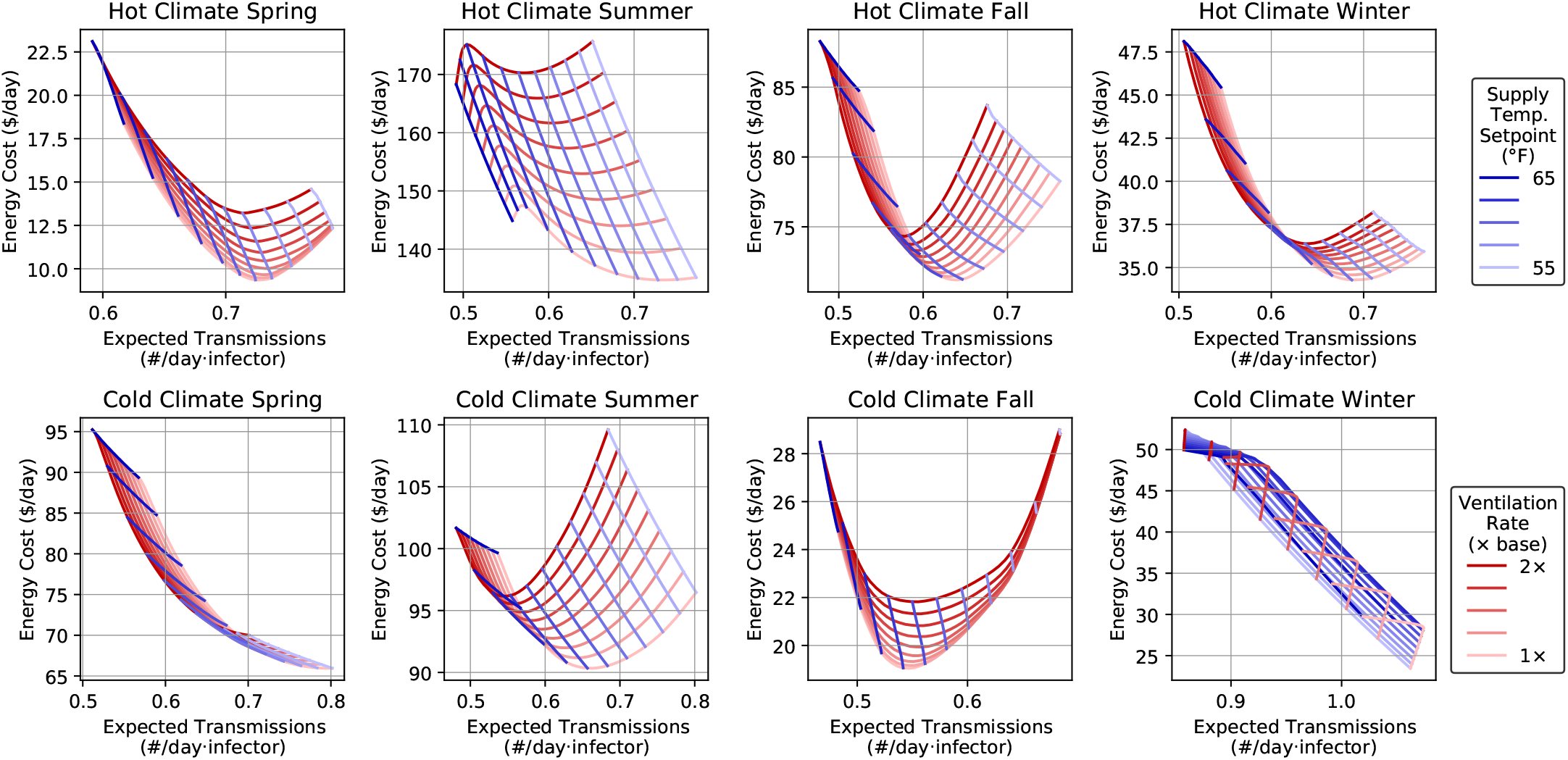
Example optimization tradeoffs for different seasons and cities. Plots show how the two-dimension optimization range gets mapped onto the to primary objective functions. Red lines hold ventilation rate constant and vary supply temperature setpoint, while blue lines hold supply temperature constant and vary ventilation rate.

**Figure 8:**
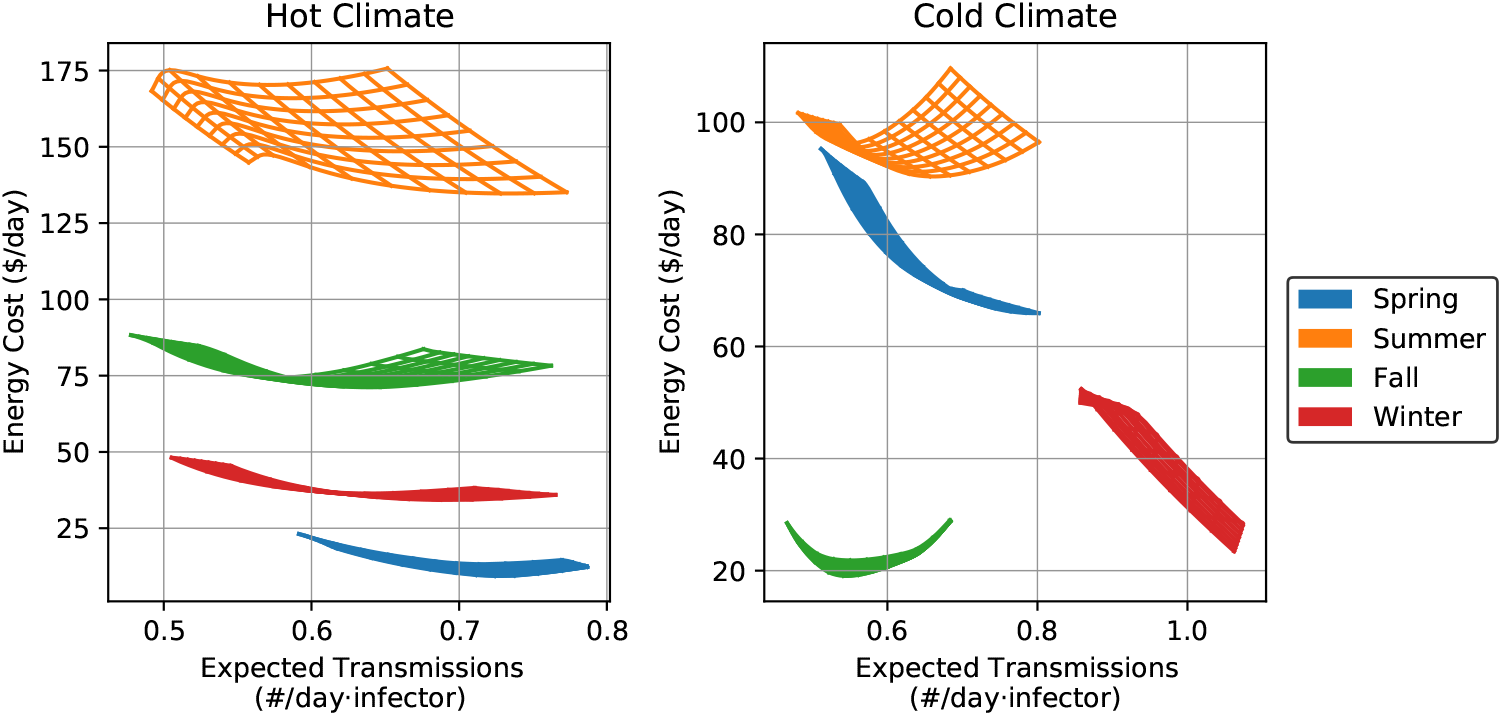
Example optimization tradeoffs for different seasons and cities. Plots show the same values as in Fig. 7 but with all seasons plotted together for each climate.

Before moving on, we highlight some of the key features of these mappings. First, for the cases that come to a sharp point in the upper left corner, it is evident that the value of the minimum ventilation rate becomes nearly degenerate at high values of the supply temperature setpoint. The explanation for this behavior is that the higher supply temperatures result in increased *total* supply airflow, and so the economizer will naturally increase outdoor-air flow (to maintain roughly a constant outdoor-air fraction). Thus, increasing the *minimum* ventilation rate essentially has no effect because the economizer is already providing more than that amount of ventilation. Second, we note that increasing the ventilation rate uniformly leads to an increase in energy use. As observed in the previous examples, this behavior is due to the aggressive economizer on this building: any time extra ventilation is energetically favorable, it will be supplied automatically by the economizer; thus, any increased ventilation that results from changing the minimum setpoint is necessarily at an energy penalty. We note that the use of energy-recovery ventilation systems could significantly reduce this cost [45], but installation of such devices would be costly and perhaps not worthwhile solely for reducing infection risk. Third, we see that increasing the supply temperature setpoint can decrease energy cost at first but then eventually begins to increase it. This is generally due to the tradeoff between latent cooling load (which decreases at higher supply temperatures) and fan power (which increases at higher flows caused by higher supply temperatures). Because both of these effects are nonlinear, their resulting sum can take on the wide variety of shapes observed in the figure depending on ambient and internal conditions. Finally, we note that the unique behavior observed in the Cold Climate Winter case is due to the fact that the system is operating at minimum-flow conditions. Thus, increasing the supply temperature setpoint above its minimum bound initially does not change the flow rate (since it is still at minimum) but requires additional energy consumption to heat the air to that temperature. Only after a sufficiently high supply setpoint does airflow rise above its minimum bound and we see more typical behavior.

Overall, we see in these example cases that the Pareto-optimal set generally but not always contains points with the lowest ventilation rate and sometimes points with the highest supply temperature setpoint. Thus, as priorities shift from energy minimization (e.g., during non-pandemic periods) to infection reduction (e.g., during a pandemic), the optimal policy is to first increase supply temperature setpoint and maybe increase the minimum ventilation rate after that. We note from the relative flatness of the sets in Fig. 8, that many ambient conditions make it possible to achieve significant reduction in expected transmissions with only minor increase in energy cost, and thus for a properly optimized system, energy consumption can remain nearly constant across pandemic and non-pandemic conditions. However, for systems with different HVAC configurations, different behavior might be optimal, and so it is important to consider the specifics of each application.

#### 4.2.3. Optimization Accuracy

To close the examples, we briefly discuss the accuracy of the proposed framework with respect to whether the optimized actions deliver the benefits predicted by the models. For this purpose, we choose representative weeks for each climate and season (corresponding to the eight cases in Fig. 7) and solve the Pareto optimization for each weekday. Note that this Pareto search accounts for thermal comfort constraints, and so on some days, there may be very few feasible points. From the set of Pareto-optimal solutions, we choose three representative points: the point that minimizes energy cost, the point that minimizes infection rate, and a point midway between them. For each of those points, we then run a new EnergyPlus simulation using the corresponding setpoints to see what the BMS data would be for those setpoints. We then calculate metrics from that data and compare to the metrics predicted by the models in the optimization problem. We also include values for the default operation of the building with both decision variables at their lower bounds.

A key challenge with this approach is that due to the simplistic disturbance forecast used (which essentially just reuses the disturbances from one week ago), it is likely that the model predictions may be systematically off due to week-to-week changes in disturbances (primarily due to solar radiation and heat exchange with the ambient). However, because the model predictions are being used for optimization, any linear re-scaling does not affect the location of the optimal solutions. Thus, to avoid the poor disturbance forecast influencing accuracy metrics, we re-scale the model’s predictions to match the actual data using one single scale factor for the entire week. This procedure will correct for the effect of the disturbance schedule, but any other model deficiencies will still appear in the plots. These results are shown in Figs. 9 and 10 for the Cold and Hot climates respectively.

**Figure 9:**
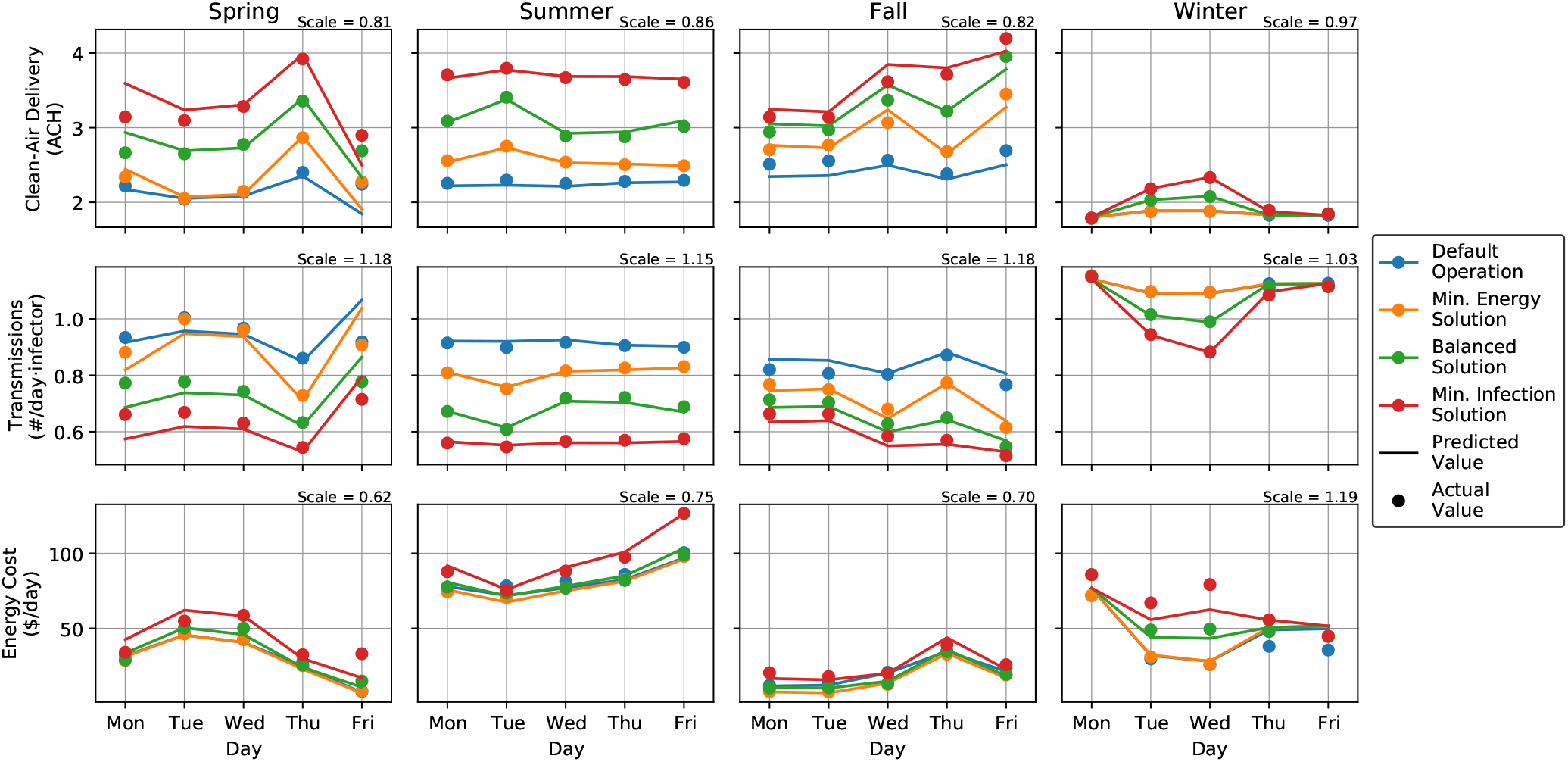
Predicted and actual optimization metrics for the Cold climate. Colors correspond to different representative points on the Pareto surface. Actual values are taken from EnergyPlus simulations using the optimized decision variables, while predicted values are metrics predicted by the dynamic models (and scaled by the indicated scale factors).

**Figure 10:**
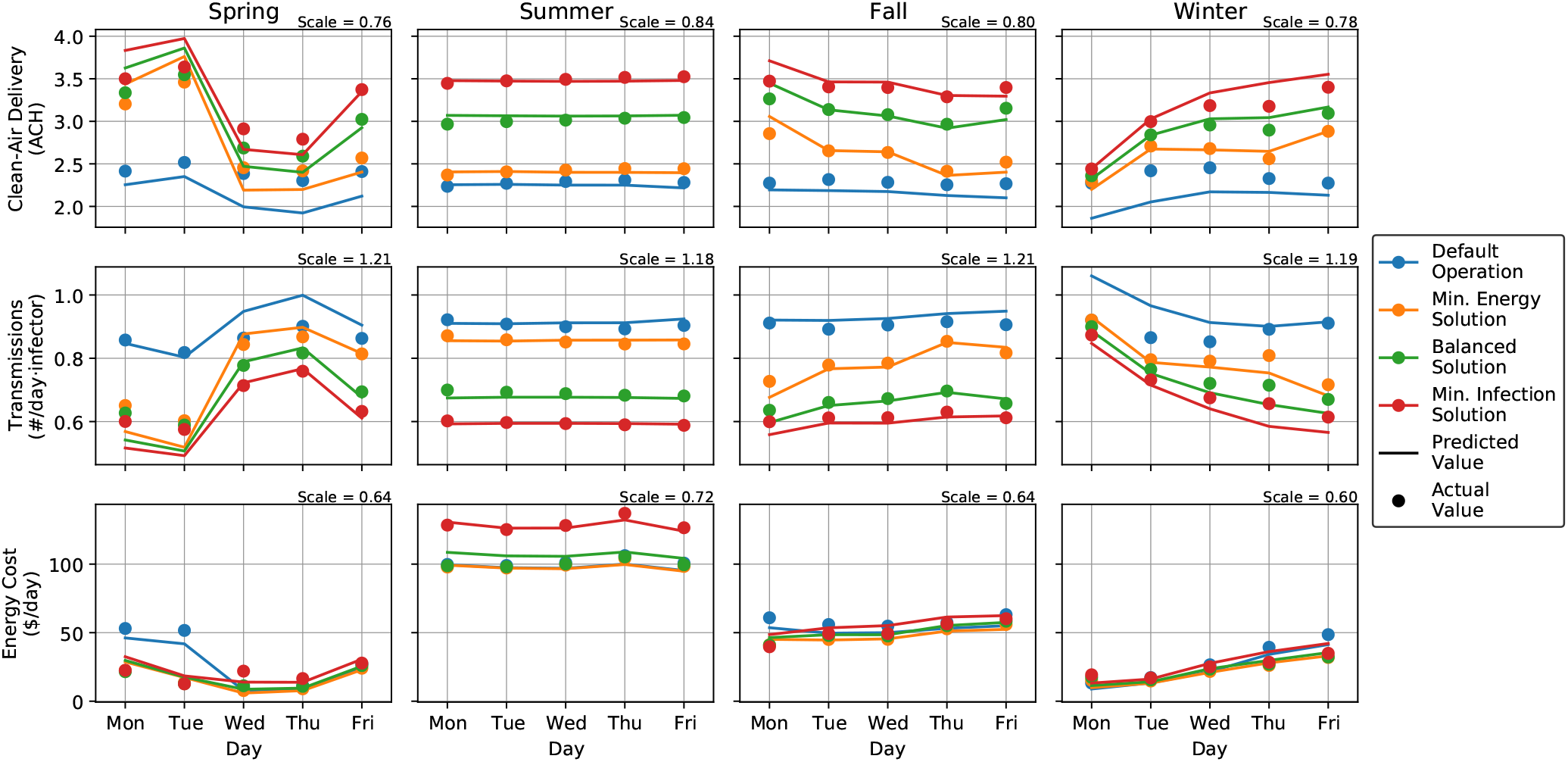
Predicted and actual optimization metrics for the Hot climate. Plots are as in Fig. 9.

From these results, we see that after correcting for the poor disturbance forecast, the models accurately predict the resulting metrics for each representative solution. Although there are some cases where accuracy is not satisfactory (in particular, the winter energy cost in Fig. 9 and the spring clean-air delivery in Fig. 10), the dynamic models do capture the available operating space for the chosen representative solutions. We note in particular that the Balanced solution often achieves noticeable reduction in infection rate with minimal (if any) impact on energy consumption. Furthermore, because the scale correction does not affect which solutions are Pareto-optimal, even the low-quality disturbance forecast used in this example would be suitable for online optimization. Thus, we see that despite the relative simplicity of the proposed models, they can nevertheless capture the key behavior of real buildings and ultimately lead to actionable insights for building operators.

## 5. Conclusions and Future Directions

In this paper, we have presented a modeling and multiobjective optimization framework to assess the tradeoff between airborne disease transmission risk and energy consumption associated with the operation of building HVAC systems. Using a physics-based dynamic model for the hypothetical concentration of airborne infectious particles that would be produced by an infector in the space, the risk of airborne transmission can be estimated via the expected exposure of susceptible occupants to those particles. By accounting for filtration, ventilation, and other infectious-particle removal sources, this model can be used with real building data and space parameters to assess the infection risk in buildings as operated. Furthermore, by coupling to additional models for space temperature and humidity as well as the action of the HVAC regulatory control system, it is possible to predict the airflow that will be delivered to the space, from which infection risk and energy consumption can be estimated. This overall model can then be embedded within a model-based optimization routine to determine Pareto-optimal values for operational setpoints and design variables. These optimization results thus provide quantitative guidance to building managers who can then make decisions consistent with their current health and sustainability goals.

To illustrate the insights provided by the proposed framework, we have shown a series of examples that apply the proposed methodology to real and simulated building data. Using BMS measurements from a real office space and gym, we show how clean-air delivery and infection rates can be estimated for each space. We then show that the proposed models can predict the future behavior of the HVAC system with sufficient accuracy to be used for optimization purposes. Finally, we have shown computed Pareto sets for the supply temperature setpoint and activation of supplemental in-zone filtration devices, illustrating that both variables can reduce infection risk but unfortunately require additional energy consumption. Using EnergyPlus simulations, we have shown how infection risk and energy consumption vary throughout the year for a typical office building for different values of the supply temperature setpoint and minimum ventilation rate. We have then presented example Pareto tradeoff curves showing that increasing the supply temperature setpoint is often the most energy-efficient way to reduce infection risk, but also that the associated energy penalties vary significantly with weather conditions. We have closed by validating the proposed models’ predictions for both baseline and optimized operating setpoints. Together, these examples illustrate that although the proposed models are relatively simple, they nevertheless capture the key building trends and can thus be a valuable source of insight for building operators.

In the future, we hope to extend the proposed framework to incorporate new indoor-air quality data sources (e.g., CO_2_ and PM sensors) to generalize optimization goals. For example, the excess CO_2_ concentration from occupants’ respiration can be correlated with airborne disease transmission risk [38, 42, 64] and thus provides a rich additional datastream to calibrate the model and improve the optimization accuracy. Even when airborne disease transmission is no longer a major concern, the average CO_2_ concentration could be optimized instead, with the primary tradeoff being between the increased energy consumption associated with ventilation and the wellness benefits to occupants associated with better indoor-air quality [45]. In this context, new design and operational variables could be introduced (e.g., whether to install energy-recovery ventilation and whether to activate it on a certain day) and optimized to account for specific space and climate characteristics. Such functionality would help support a paradigm shift to combat indoor respiratory infection and improve occupant wellness in the aftermath of the COVID-19 pandemic [5]. Overall, we hope that model-based optimization will increasingly be used as a tool to guide design and operation of building HVAC systems to improve health and sustainability.

## Data Availability

All data is included in the manuscript or referenced works.

## Acknowledgements

This research did not receive any specific grant from funding agencies in the public, commercial, or not-for-profit sectors. Real building datasets were provided by Johnson Controls International. The authors would like to thank Michael Wenzel, Mohammad ElBsat, Anas Alanqar, and Chenlu Zhang for their helpful discussions.

## Appendix A. Infection Model Discussion

### A.1. Clean-Air Delivery Calculation

To model the time variation in infection quanta concentrations, we note that infectious particles are released into the air by the infectious individual and then removed by a variety of mechanisms. To standardize modeling of these effects, we define the concept of “clean-air delivery” to the space as follows: for a piece of equipment or physical process *j* that deactivates or captures infectious particles at a total rate *r*_*j*_ (measured in units of q/s) from an air stream with concentration *C* (measured in units of q/m^3^), the corresponding clean-air delivery rate is defined as

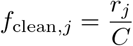

which thus has units m^3^/s corresponding to a hypothetical volumetric flow (and can be converted to air-change rates by dividing by the space volume). We use this definition because for most disinfection processes, *r*_*j*_ is directly proportional to *C*, and thus *f*_clean,*j*_ can be expressed as a simple fraction of airflow through the device. In addition, this formulation provides a common basis for comparison of different disinfection devices: multiple devices providing the same clean-air delivery rate thus become fungible from an infection-risk perspective and can be assessed in terms of energy consumption or other considerations to choose the best among them.

One complication of this clean-air framework is that for devices and processes that operate at the particulate level (i.e., deposition and filtration), the removal rate *r*_*j*_ depends on the size distribution of the particles containing the infectious material [4]. When the inlet size distribution is known, accounting for size-dependent effects is straight-forward for a single device. However, because their action leads to changes in the airborne size distribution, it is not longer possible to fully and exactly describe the clean-air characteristics of a device without knowing (and accounting for the inherent recirculation effects on) the size distribution of airborne particles. In the interest of simplicity, we assume a constant steady-state particle size distribution (using the size bins defined by ASHRAE standards) and calculate removal rates accordingly. From previous studies for influenza, Azimi and Stephens [37] states that 20% of viral material is found within E1 particles (diameters 0.3 to 1 µm), 29% within E2 particles (diameters between 1 and 3 µm), and the remaining 51% within E3 particles (diameters between 3 and 10 µm). Assuming these values hold for the pathogens of interest, an effective clean air delivery rate can be calculated via a weighted average of the removal rates for a representative particle diameter within each size bin. However, see discussion in Section A.5 for how this effect could be modeled more accurately without significant changes to the existing framework.

Aside from size-dependent effects, the calculation of the clean-air delivery rate for most processes and devices is straightforward. For example, airborne particles naturally deposit on the ground at a given velocity *ν*_deposition_ due to gravity. Thus, in a room with volume *V* and height *h*, the removal rate of such particles by deposition is given by

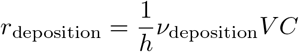

Similarly, due to natural decay of the viruses (assumed to occur with a first-order rate constant *k*_decay_), the removal rate due to natural decay is thus given by

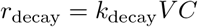

In practice, this decay rate may vary with temperature and humidity within the space [4], but for simplicity here we assume it is truly constant. As modeled above, we combine these two effects as “natural” disinfection sources in Eq. (1) with

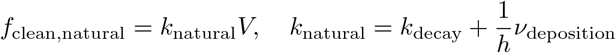

Note that they scale with the total *volume* of the space and occur independently of any HVAC operational decisions.

For standalone filtration devices, the clean-air calculation is also straightforward: assuming a total volumetric flow *f*_filter_ through the filter, which captures a fraction *η*_filter_ of the particles the total removal rate is given by

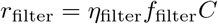

and so the clean-air delivery is simply

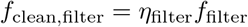

Effective filtration efficiencies for various filter grades are known [44], modulo uncertainty in size dependence. Similarly, for UV devices that kill a given fraction *η*_UV_ of the microbes passing through them, the clean air production is

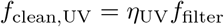

If properly designed, UV disinfection tunnels can achieve very high disinfection rates *η*_uv_ ≈ 1 [28], but the performance of smaller standalone devices may vary.

For devices operating in parallel, their total clean-air delivery is a simple sum of their individual values. Since all auxiliary in-zone devices are effectively in parallel, we thus include the summation term in Eq. (1), noting that the *maximum* clean-air capacity 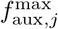 of a given auxiliary device needs to be scaled by the extent *a*_aux,*j*_ to which it is currently active. Note that for some devices *a*_aux,*j*_ may vary continuously, while for others, it may take only a binary value indicating whether the device is on or off.

For devices operating directly in series, however, the total clean-air delivery must account for the fact that the clean-air delivery for each device must be calculated in terms of the original inlet concentration rather than that device’s particular inlet concentration. The canonical example of this configuration is in the AHU: when air is returned to the AHU, a given fraction *x*_*a*_ is vented to the ambient (and replaced with a corresponding volume of outdoor air), after which it passes through a filter with disinfection efficiency *η*_filter_ and possibly also through a UV kill tunnel that kills a fraction *a*_uv_*η*_UV_ of microbes in the airstream. Taken individually, the clean-air rates of each device would be given as

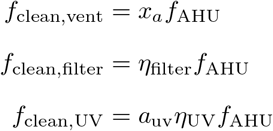

However, because these devices operate in series, we must account for the fact that each device processes the outlet of the previous device. Thus, the inlet concentration to the filter is only 1 − *x*_*a*_ times its initial value, and thus the filter provides only (1 − *x*_*a*_)*η*_filter_ fractional disinfection. The overall clean-air delivery for the AHU is thus given by

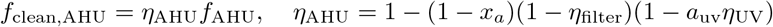

consistent with the definition in Eq. (1). We can see immediately from the functional form that *f*_clean,AHU_ ≤ *f*_AHU_ which makes intuitive sense: the AHU cannot clean more air than passes through it.

### A.2. Infection Probability and Received Dose

To estimate the risk of infection in a zone, we calculate a hypothetical number of transmissions during the course of the evaluation time period (commonly on a daily basis) as given by Eq. (2). Desired limits can be placed on this value to guide operational and HVAC decisions. The key question then becomes how the time-varying trajectory of *C* resulting from the hypothetical infectors leads to infection risk for susceptible occupants in the space.

In the ideal case, infection risk would be calculated on an individual basis and then aggregated across the building to determine overall infection risk. Using the dynamic model Eq. (1) for the airborne infectious quanta concentration *C* in each zone, we can thus estimate an individual’s received dose throughout the day and ultimately compute that individual’s infection probability assuming a constant hazard rate (expressed in units of q^−1^) for new infection. For a susceptible individual *s*, we have

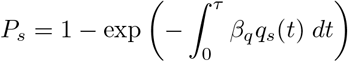

with *P*_*s*_ giving that individual’s infection probability in terms of the individual’s infectious dose rate *q*_*s*_, which is itself given by

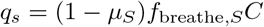

We note that at steady-state conditions (i.e., *dC/dt* = 0), the model in Eq. (1) gives *C* = *q*_*I*_*/f*_clean_. Substituting these values (and converting the time-varying integral to a simple multiplication) gives

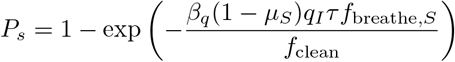

which is equivalent to the common steady-state presentation of the Wells-Riley equation [4, 37, 40]. However, in variable-volume HVAC systems, the steady-state assumption is rarely satisfied, as the regulatory control layer constantly adjusts supply airflow *f*_AHU_ to maintain temperature within the space in response to variation in weather and occupancy. Thus, the resulting quanta concentration fluctuates throughout the day and should be modeled as a time-varying quantity to more accurately represent infection risk.

After each individual’s infection probability has been calculated, these values can then be aggregated over all building occupants to find the average number of transmissions given by

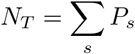

with distributional information available if needed. This value can of course be converted to a *rate* by dividing by the total evaluation time *τ*. Unfortunately, this person-based approach is not practical for implementation in real buildings. First, it implicitly requires that the infection quanta concentration be modeled in every space within the building, which may not be worthwhile for low-occupancy or otherwise low-risk zones. Second, even if every space were modeled, it is generally not possible to track each individual’s movement throughout the building. Third, since we are interested in the HVAC operation of the zones, it is more practical to adopt a zone-based approach in which infection risk is associated with spaces rather than individuals.

To address these limitations, we take an approach similar to Bazant and Bush [4] and make a linear approximation as follows:

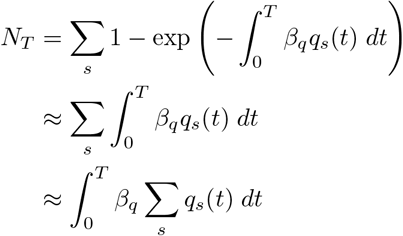

Noting that Eq. (1) has already defined *q*_*S*_ = ∑_*s*_ *q*_*s*_(*t*) in the model, substituting this value gives the equation for *N*_*T*_ presented in Eq. (1). The main benefit of this formulation is that we can now split the whole-building *R* on a zone-by-zone basis. Thus, separate actions can be taken in each zone based on that zone’s contribution to overall infection risk.

In arriving at this formulation, we have made use of the fact that 1 − exp(−*x*) ≈ *x* for small values of *x*, which allows us to remove the nonlinearity and shift the summation inside the integral. We note also that the linear approximation is conservative, i.e., uniformly *over-estimating N*_*T*_, due to concavity of the exponential function. Assuming that the infection risk is fairly well controlled, buildings should be operating in the range where this approximation is highly accurate, at least over the course of one day. Expressed differently, if we are relying on the nonlinearity of the infection probability curve as a source of infection risk reduction, then we have already failed at infection control. Thus, the linear approximation is always reasonable when modeling the mitigation of airborne transmission risk.

### A.3. Key Parameters

The infection quanta model and infection rate calculation Eqs. (1) and (2) include a number of key parameters related to infector and occupant behavior. Specifically, these quantities are the the breathing rates *f*_breathe,*I*_ and *f*_breathe,*S*_ of infectors and susceptibles, the mask effectivenesses *µ*_*I*_ and *µ*_*S*_, and finally the infector exhaled quanta concentration *N*_*I*_. We discuss these parameters in more detail as follows.

The breathing-rate parameters *f*_breathe_ are relatively straightforward to estimate, although their values are somewhat uncertain. The EPA provides values [63, Table 6-2] for average breathing rates based on age and activity level. For a 40-year-old individual, these values range from 0.276 m^3^/hr for sedentary activity to 2.94 m/hr for high-intensity activity. For typical office or commercial environments, a default value of 0.67 m^3^/hr is reasonable, but this value should be adjusted for specific spaces with different activity levels (e.g., gyms) and possibly also for different age distributions (e.g., in an elementary school). Note that because we generally do not know which specific individuals in the space are infectious, we will almost always assume *f*_breathe,*I*_ ≈ *f*_breathe,*S*_. However, the values can be considered separately in edge cases where distinct separate values are known.

For the mask effectiveness parameters *µ*_*I*_ and *µ*_*S*_, unfortunately there is very large uncertainty relating to both mask material and fit. For example, an N95 mask may have a theoretical filtration efficiency above 99%, but if worn incorrectly (so that a significant amount of air can escape around the edges of the mask), then its effective efficiency may be significantly lower. Given this variability, we use a default value of roughly 70% filtration efficiency. As with the breathing-rate parameters, we generally do not know which specific individuals are infectious, and so there may not seem to be clear motivation to use different values for *µ*_*I*_ and *µ*_*S*_. However, when aerosols are exhaled by humans, they generally contain a large amount of water that will evaporate as the particles come to equilibrium with the humidity in the space. As this evaporation occurs, the particles shrink in diameter by roughly a factor of 2 at 50% relative humidity [4]. Thus, we generally expect that *µ*_*I*_ *> µ*_*S*_ since the particles are bigger when exhaled through the infector’s mask than when they are inhaled through the susceptibles’ masks. Although this effect is challenging to quantify, we retain separate *µ*_*I*_ and *µ*_*S*_ in case accurate experimental values are obtained in the future.

Finally, for the infector’s exhaled quanta concentration *C*_*I*_, we note that this value is commonly characterized in conjunction with the breathing rate *f*_breathe,*I*_ as a single parameter *q* (denoted *q*_*I*_ in our equations above) giving a quanta generation rate in quanta per time [37, 40]. However, given that COVID-19 is transmitted primarily in respiratory aerosols, it is suggested by Bazant and Bush [4] that the value be disaggregated as separate *C*_*I*_ and *f*_breathe,*I*_ as we have modeled it, with *C*_*I*_ ranging from 10 q/m^3^ for quiet breathing, to q/m^3^ while speaking, and as high as 1000 q/m^3^ while singing. This choice of parameterization makes it clear that the dependence of infection risk (as determined by the total received quanta dose) is roughly *quadratic* in the average breathing rate. Of course, the true generation rate of infectious quanta is complicated by additional factors (e.g., intermittant coughing and variation in viral load due to natural disease progression), but this parameterization at least captures the fundamental trends with respect to activity level and vocalization.

### A.4. Susceptible and Infectious Occupants

Two of the most important inputs in Eq. (1) are the numbers of infectious and susceptible occupants *N*_*I*_ and *N*_*S*_. Given that these values cannot be directly measured, we must *choose* appropriate values based on typical occupancy profiles and other considerations.

For the infectors *N*_*I*_, one option is to choose the value based on the incidence of disease in the general population. Such statistics are generally available from the CDC and other organizations and can be multiplied by the design occupancy of a zone. Unfortunately, these values can vary significantly with time and location, which makes them difficult to incorporate into quantitative analysis. Thus, we instead opt for a conditional approach as follows: rather than trying to estimate the *actual* number of infectors *N*_*I*_ in the space, we instead *assume* that *N*_*I*_ = 1 and thus calculate infection risk conditioned on the fact that that one infectious individual has made it into the space. (Note that to account for the typical occupancy cycles, we do not assume a constant *N*_*I*_ ≡ 1, but rather set *N*_*I*_ = 1 only during occupied hours and leave *N*_*I*_ = 0 during unoccupied hours.) With this assumption, the infection rate *N*_*T*_ from Eq. (2) thus scales linearly in the number of infectors (i.e., 2 infectors in the space will on average cause 2*N*_*T*_ infections over the course of a day, 3 will cause 3*N*_*T*_, etc.). We know from the mathematics that this approach will *overestimate* the number of new infections when the number of infectors is larger (since the susceptible population is correspondingly reduced), but if the situation is already that dire, HVAC measures alone will not be sufficient to prevent disease transmission, and the prudent choice is to simply close the building. The main point is that the *N*_*I*_ = 1 assumption decouples our simulations from external conditions while still providing valuable insight into the inherent risk posed by each zone. In addition, dividing the number of transmissions *N*_*T*_ by the average number of infectors *N*_*I*_ = 1 thus gives an estimate of the *reproductive number* for the space, i.e., the number of additional transmissions caused by each infector. To avoid runaway transmission of disease, buildings should thus be operated so that *R* ≤ 1 (although due to uncertainty in other parameters, a considerable safety margin is likely warranted).

Finally, for the number of susceptibles *N*_*S*_, the main consideration is to account for possible immunity in some or all building occupants. The conservative approach is of course to assume no occupants are immune, and thus *N*_*S*_ is simply the time-varying number of occupants in the space (less the chosen number of infectors *N*_*I*_). Where vaccination rates are known, *N*_*S*_ can be further reduced by the fraction of occupants who are vaccinated, perhaps attenuating slightly to account for the fact that immunity is not absolute. For buildings where full time-varying occupancy profiles are not known, representative profiles are available in ASHRAE Standard 90.1 [55] that can be scaled by the design occupancy for each zone. Alternatively, time-varying CO_2_ measurements can also be used to estimate occupancy patterns [65–67].

### A.5. Size-Resolved Infection Model

As alluded to in Section A.1, the size dependence of many infectious particle removal mechanisms means that the size distribution of airborne particles will vary over time based on the size-dependent balance of particle generation (by infectious occupants) and removal (via clean air delivery). The approach proposed previously is essentially to assume that the size distribution of infectious particles within the zone is equal to the size distribution as exhaled by infectious occupants. Unfortunately, this assumption may not hold if clean-air sources with strongly size-dependent efficiency are used. To more accurately model this effect, the best modification is to simulate a separate instance of Eq. (1) for a representative set of particle diameters. For example, using the ASHRAE-defined size bins, one would simulate separate models for *C*^E1^, *C*^E2^, and *C*^E3^ giving the infectious particle concentration (still measured in q/m^3^) only for particles within the E1, E2, and E3 size bins. All model parameters that exhibit size dependence (including *k*_natural_, 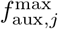, *η*_filter_, *µ*_*I*_, *µ*_*S*_, and *C*_*I*_) can then take on their appropriate values for particles within that range.

To calculate the expected transmission rate in this size-resolved model, the only modification is that exposure should now aggregate over the various size bins. Specifically, Eq. (2) should be modified to calculate

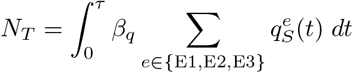

in which the 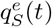 are the size-resolved exposure rates calculated from the separate simulations of Eq. (1) for each size bin *e*. In the interest of simplicity, we have chosen not to adopt this model for the results presented in this paper, but its application is relatively straightforward provided that the number of size bins is kept reasonably small.

## Appendix B. Thermal and Energy Model Discussion

### B.1. Temperature and Humidity Models

In addition to the net advective transfer provided by the supply and return air streams, the temperature model in Eq. (3) considers conductive heat transfer with the ambient proportional to the temperature difference *k*_*az*_(*T*_*a*_ − *T*_*z*_), and all other sources of heat gain (e.g., plug, lighting, and metabolic loads), are assumed to be lumped into the *Q*_*z*_ input. For simplicity, we do not explicitly model the temperature dynamics of solid-mass component (as operating temperatures are relatively constant, and thus temperature-dependence of this effect can be largely neglected), and we do not explicitly calculate the effect of solar radiation (as accurate solar forecasts are often not available). However, these effects can be added to the model where relevant data is available. Separate zones within a building are assumed to be physically thermally isolated, and so no exchange (mass or energy) is modeled. Direct infiltration from the ambient is not considered, while exfiltration is implicitly included in the return-air stream. For zones that require heating (e.g., exterior zones in cold climates), we include the *Q*_*h*_ term as a direct addition of heat to remain agnostic to the heating source. This supplemental heating may be supplied by reheat coils inside the VAV boxes or by in-zone electric heaters. In any case, since it operates at the room level, we can accurately model it as a direct addition of heat.

As with the temperature model, the humidity model in Eq. (3) considers advective transfer from the supply air and an internal generation term (due primarily to occupants). However, since infiltration from the ambient is not considered, there is no direct exchange term between *ω*_*z*_ and *ω*_*a*_. For robustness, the value of *ω*_*z*_ could be checked (and clipped appropriately) to ensure that it does not exceed the saturation point *ω*_sat_(*T*_*z*_). However, since such conditions should only arise during unrealistic operating scenarios (e.g., if the cooling coil is disabled during hot and humid ambient conditions), this check can generally be skipped. In the interest of brevity, we do not consider humidification equipment in the current model. Where present and active, it would be added as an additional generation term in the moisture model for in-zone equipment or in the AHU coil model for AHU equipment.

### B.2. AHU Coil Model

We assume that the AHU controls its supply air temperature to setpoint using a heating coil and a cooling coil in sequence. Air passing through the AHU is thus heated or cooled depending on whether its temperature is below or above the setpoint temperature 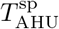. The previous Eq. (4) accounts for the action of the appropriate coil. In the interest of simplicity, we present these equations assuming unbounded cooling capacities (which means the setpoint can always be met), as total loads are implicitly constrained by bounds on flow. However, extension to directly account for finite coil capacity is straightforward.

To start, we calculate the AHU inlet conditions by mixing the outdoor-air and recirculating air streams. A straightforward energy balance gives

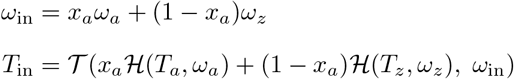

in which *ℋ* (*T, ω*) gives the specific enthalpy of an air stream at temperature *T* and humidity *ω*, while *𝒯* (*h, ω*) gives the temperature of an air stream at specific enthalpy *h* and humidity *ω*. From here, if 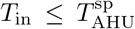, the air is heated directly to the setpoint with no change in humidity. Thus, the AHU outlet conditions are 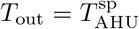 and *ω*_out_ = *ω*_in_. Alternatively, if 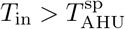, the air is cooled as discussed next.

Because of the possibility of condensation, the cooling coil requires some additional modeling to determine the outlet humidity. For this purpose, we follow the common “contact mixture analogy” (e.g., as in Seem and House [59]). With this formulation, we assume that a fixed fraction of air (*x*_bypass_) bypasses the cooling coil completely, while the remaining fraction (1 − *x*_bypass_) equilibrates with the coil. As before, we assume that the cooling coil meets the setpoint, which gives 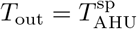. From this value, we can back-calculate the coil-equilibrating conditions as

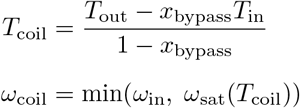

in which *ω*_sat_(*T*) gives the saturation humidity at temperature *T*. This term is responsible for modeling the dehumidification provided by the coil, which is often a significant fraction of energy consumption in humid climates. Note that the first equation assumes linear mixing for the recombination of coil and bypass streams. We calculate the outlet humidity as

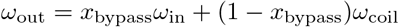

which will always be less than or equal to the inlet humidity *ω*_in_.

Summarizing the heating and cooling models, we thus have the following relationships:

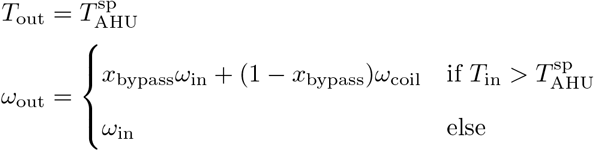

From these conditions, we can thus calculate the thermal loads on the coils as follows:

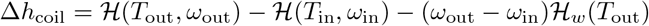

in which *ℋ* (*T, ω*) is the specific enthalpy of wet air at temperature as before, and *ℋ*_*w*_(*T*) is the specific enthalpy of liquid water. Note that this expression is written so that it gives the heat gained by the air stream, with a positive value indicating the stream was heated and a negative value indicating the stream was cooled. We can thus split into individual heating and cooling components as

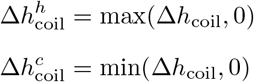

The calculated *T*_out_, *ω*_out_, 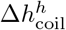, and 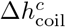 then become the AHU conditions *T*_AHU_, *ω*_AHU_, 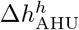, and 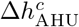 indicated in Eq. (4). As above, we do not consider humidification equipment, but where present, it could be accounted for by increasing *ω*_AHU_ up to a given setpoint.

### B.3. Fan Power Model

The fan power model in Eq. (5) defines electricity consumption of the fan as a cubic function of airflow *f*. Physically, the energy consumed by the fan is given by

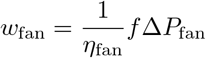

where *η*_fan_ is the mechanical efficiency of the fan, and Δ*P*_fan_ is the pressure rise across the fan. The cubic dependence on *f* comes from the fact that the pressure rise of the fan is equal to the pressure drop through the ductwork, which is commonly modeled as proportional to the square of flow. These considerations suggest that *w*_fan_ ∝ *f* ^3^. In practice, the mechanical efficiency of the fan will vary over its operating range, and thus the cubic term alone is not sufficient to match the actual performance of the fan. By adding the additional linear and quadratic terms, this variation can be captured. Note that some sources will suggest adding an additional constant term to better match experimental behavior. However, we require for our application that *w*_fan_ is zero when *f* is zero (e.g., during unoccupied hours when the HVAC system is shut off), and so we do not include this term to avoid artificial inflation of predicted electricity consumption.

The most accurate way to determine the coefficients *c*_*i*_ in the fan model is to obtain a dataset of (*f, w*_fan_) pairs from the actual fan and use standard regression. Unfortunately, it is uncommon to directly measure *w*_fan_ is real buildings, and so this approach is not practical. Instead, we define one set of coefficients 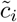 to match a representative AHU fan with efficiency 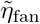 and pressure rise 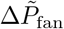. We then rescale the 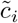 linearly based on the design values for *η*_fan_ and Δ*P*_fan_ (which are generally readily available) for the each AHU we model. This approach gives reasonable accuracy for a variety of zones and buildings.

A key requirement for the fan model is that it be able to account for the different electricity consumption associated with different filter types. In general, higher-efficiency filters require higher pressures to deliver the same airflow. This additional pressure must be overcome by the fan, which in turn increases electricity consumption. To model this effect, we simply rescale the coefficients to match the expected fan power at maximum flow *f* ^max^. From manufacturer’s data, we know the pressure drop Δ*P*_filter_ associated with each filter type. We then assume that the design fan pressure rise Δ*P*_fan_ corresponds to a baseline filter type with pressure drop 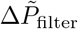. Combined with the scaling for design pressure drop, we thus have the model

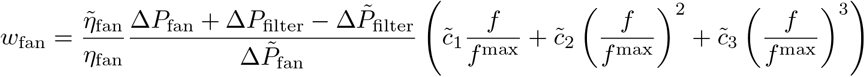

Merging the prefactors with the representative 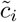 coefficients thus gives the specific *c*_*i*_ coefficients consistent with Eq. (5).

## Appendix C. Controller Model Discussion

### C.1. Zone Temperature Control

As shown above in Eq. (7) the controller model chooses values of *f*_AHU_ and *Q*_*h*_ by clipping the hypothetical values 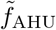 and 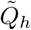 that would move zone temperature to the respective cooling and heating bounds 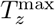 and 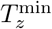. Based on system structure, it is natural to first calculate *f*_AHU_ (as it has a nonzero lower bound due to the minimum outdoor-air ventilation constraint) assuming *Q*_*h*_ = 0 and then calculate *Q*_*h*_ second to provide the necessary supplemental heating. The key question is how to calculate the hypothetical values 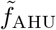 and 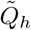.

The ideal strategy to calculate the hypothetical variables is to implicitly solve (the discretization) of the thermal ODE model from Eq. (3). Forward simulation of this model defines the successor value of *T*_*z*_ as a function of the current *T*_*z*_, the chosen *f*_AHU_ or *Q*_*h*_, and the values of the remaining model inputs. Thus, we can solve those equations for the 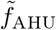 and 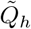 that drive *T*_*z*_ to 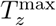 and 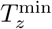 respectively, i.e., solving

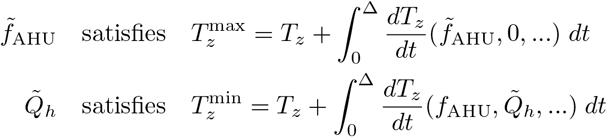

in which the “…” represent the remaining model inputs (which would not all be known in a real system but are known to the model). Note that for physical reasons, both variables must be nonnegative, but since that restriction is enforced by clipping after the fact, we place no sign restrictions on 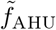 and 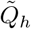. From the model structure, we know that a suitable value 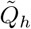 always exists and a suitable 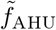 exists under standard operating conditions (i.e., *T*_AHU_ *< T*_*z*_).

Unfortunately, because *f*_AHU_ enters the ODE in a product with the state variable *T*_*z*_, an exact solution to the first equation requires iterative methods. The use of such algorithms can lead to slow and inconsistent solution times, and so we would like to avoid them. Thus, we instead opt for an approximate solution that is non-iterative. To determine 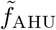 we assume that the *ρ*_*a*_*c*_*a*_*f*_AHU_(*T*_AHU_ − *T*_*z*_) term in Eq. (3) can be approximated by a constant value *Q*_*c*_ over the simulation interval. By removing the *f*_AHU_ *× T*_*z*_ nonlinearity, our unknown variable now enters the ODE linearly and thus can be solved for directly using the exact discretization of

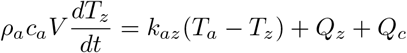

with the condition that 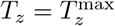 after one timestep. With the corresponding value 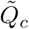, we thus back-calculate

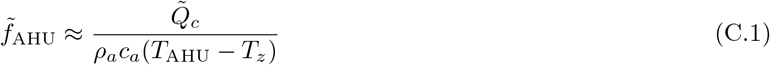

In the event that the denominator is zero, we set 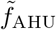 to *±*∞ depending on the calculated sign of 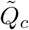. In cases where *T*_AHU_ and *T*_*z*_ are constant during the integration interval, this calculation is exact, and the accuracy deteriorates as *T*_AHU_ and/or *T*_*z*_ vary within the interval. By assumption, *T*_AHU_ will be equal to its setpoint 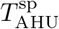, and so there is little to no approximation error there. Similarly, during occupied hours, *T*_*z*_ will generally be near the cooling setpoint 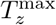 or heating setpoint 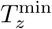 depending on the season. However, during transitions at the beginning of the occupied period, *T*_*z*_ can change very quickly as the controller tries to restore setpoint, and so it is generally a good idea to replace *T*_*z*_ with an average value 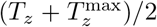 to avoid overshoot. In any case, as long as the zone temperature setpoint stays reasonably constant throughout the occupied period, this calculation is sufficient to accurately model airflow to the space.

Note that it is in Eq. (C.1) where the primary impact of the supply-temperature setpoint 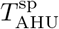 is considered. Specifically, when *T*_AHU_ is higher and thus closer in value to *T*_*z*_, we see that the denominator becomes smaller, and so the delivered airflow 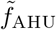 increases. A similar effect would also be possible by decreasing *T*_*z*_ (via its setpoint 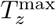), but due to the potential impact on occupant comfort, we do not directly consider that as a manipulated variable. In any case, the key observation is that the impact of *T*_AHU_ on airflow (and thus also on infectious-particle concentration) is implicit due to the action of temperature control in the space.

After clipping 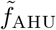 to find the true *f*_AHU_ as in Eq. (7), we can thus determine 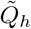 via the discretization of

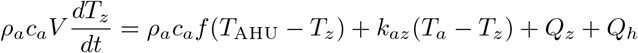

to arrive at 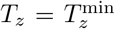. Here, we can solve for 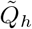 using the exact linear formulas as for 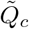 above. The resulting value is then clipped to *Q*_*h*_ as in Eq. (7).

### C.2. Outdoor Air and Disinfection Device Control

The outdoor-air fraction *x*_*a*_ is chosen in accordance with standard economizer logic: if the outdoor air temperature is colder than the pre-configured threshold 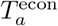 (which means that the outdoor air is a free source of cooling), then the controller should use the maximum outdoor-air fraction 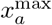; otherwise, if the air is hotter than 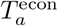 (which means the air is *not* a free source of cooling), then the controller should use the minimum 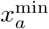. Calculation of these bounds is described next. The remaining quantities *a*_UV_ and *a*_aux,*j*_ associated with disinfection devices are simply cycled on and off in accordance with the occupancy flag *σ*.

The two outdoor-air fraction bounds 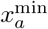 and 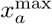 are calculated as shown in Eq. (8). For 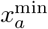, its value is simply the minimum fraction of *f*_AHU_ that will deliver the required minimum outdoor-air ventilation rate 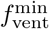 (which cycles between zero and the pre-configured bound 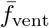 based on the occupancy flag *σ*). As discussed in the previous section, we always have that 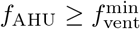, and so we know that 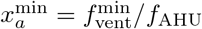 will always be between 0 and 1. For 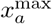, the bound is calculated as the maximum fraction of *f*_AHU_ that will keep the temperature of the resulting mixture below the supply temperature setpoint 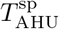 (so as to avoid an unnecessary heating load). Assuming linear mixing, this condition is exactly satisfied when

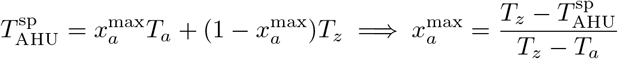

which gives a valid 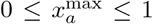 whenever 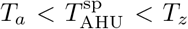. However, because those constraints are not always satisfied (in particular due to seasonal variation in *T*_*a*_), we modify the calculation to clip both temperature differences to zero, i.e.,

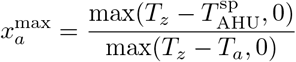

with the convention that 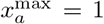 in the singular case *T*_*z*_ = *T*_*a*_. With these modifications, the calculation gives 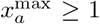 whenever 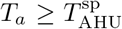 and 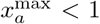 whenever 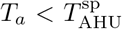 (i.e., when outdoor conditions are too cold for 100% outdoor air). This value is then clipped to be between the lower bound 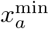 and the upper bound 1 to give the calculation in Eq. (8).

Note that the models here assume that a simple (but standard) outdoor-air economizer is responsible for outdoor-air control. However, if different equipment is present, these models may have to be modified. For example, if demand-controlled ventilation systems are present (which modulate outdoor-air flow to control indoor CO_2_ concentration to a given setpoint), then it will be necessary to model the CO_2_ concentration in the space (with an additional transient mass balance similar to the *ω*_*z*_ model) and use logic similar to the *f*_AHU_ controller model to determine *x*_*a*_. However, in the interest of simplicity, we do not consider such cases here.

## Notes

### Competing Interest Statement

The authors have declared no competing interest.

### Funding Statement

The authors received no external funding for this work.

### Author Declarations

We confirm that all ethical guidelines have been followed. IRB approval is not required for this work.

